# Region-based analysis with functional annotation identifies genes associated with cognitive function in South Asians from India

**DOI:** 10.1101/2024.01.18.24301482

**Authors:** Hasan Abu-Amara, Wei Zhao, Zheng Li, Yuk Yee Leung, Gerard D. Schellenberg, Li-San Wang, Priya Moorjani, A.B. Dey, Sharmitha Dey, Xiang Zhou, Alden L. Gross, Jinkook Lee, Sharon L.R. Kardia, Jennifer A. Smith

**Affiliations:** Department of Epidemiology, School of Public Health, University of Michigan, Ann Arbor, Michigan, United States of America; Survey Research Center, Institute for Social Research, University of Michigan, Ann Arbor, Michigan, United States of America; Department of Biostatistics, School of Public Health, University of Michigan, Ann Arbor, Michigan, United States of America; Penn Neurodegeneration Genomics Center, Department of Pathology and Laboratory Medicine, Perelman School of Medicine, University of Pennsylvania, United States of America; Department of Molecular and Cell Biology, University of California, Berkeley, United States of America; Center for Computational Biology, University of California, Berkeley, United States of America; Department of Geriatric Medicine, All India Institute of Medical Sciences, New Delhi, India; Department of Biophysics, All India Institute of Medical Sciences, New Delhi, India; Department of Epidemiology, Johns Hopkins Bloomberg School of Public Health, Johns Hopkins University, Baltimore, Maryland, United States of America; Department of Economics, University of Southern California, Los Angeles, California, United States of America

**Keywords:** Genetics, genomics, region-based analysis, cognitive function, whole genome sequencing, South Asian

## Abstract

The prevalence of dementia among South Asians across India is approximately 7.4% in those 60 years and older, yet little is known about genetic risk factors for dementia in this population. Most known risk loci for Alzheimer’s disease (AD) have been identified from studies conducted in European Ancestry (EA) but are unknown in South Asians. Using whole-genome sequence data from 2680 participants from the Diagnostic Assessment of Dementia for the Longitudinal Aging Study of India (LASI-DAD), we performed a gene-based analysis of 84 genes previously associated with AD in EA. We investigated associations with the Hindi Mental State Examination (HMSE) score and factor scores for general cognitive function and five cognitive domains. For each gene, we examined missense/loss-of-function (LoF) variants and brain-specific promoter/enhancer variants, separately, both with and without incorporating additional annotation weights (e.g., deleteriousness, conservation scores) using the variant-Set Test for Association using Annotation infoRmation (STAAR). In the missense/LoF analysis without annotation weights and controlling for age, sex, state/territory, and genetic ancestry, three genes had an association with at least one measure of cognitive function (FDR q<0.1). *APOE* was associated with four measures of cognitive function, *PICALM* was associated with HMSE score, and *TSPOAP1* was associated with executive function. The most strongly associated variants in each gene were rs429358 (*APOE* ε4), rs779406084 (*PICALM*), and rs9913145 (*TSPOAP1*). rs779406084 is a rare missense mutation that is more prevalent in LASI-DAD than in EA (minor allele frequency=0.075% vs. 0.0015%); the other two are common variants. No genes in the brain-specific promoter/enhancer analysis met criteria for significance. Results with and without annotation weights were similar. Missense/LoF variants in some genes previously associated with AD in EA are associated with measures of cognitive function in South Asians from India. Analyzing genome sequence data allows identification of potential novel causal variants enriched in South Asians.

## Introduction

Dementia is a group of neurological disorders characterized by cognitive impairment. In 2019, the estimated global economic costs of dementia was about $1.3 trillion USD [1]. The public health burden for dementia is borne disproportionately by lower- and middle-income countries, which harbor approximately 61% of affected individuals [1]. Over 50 million people worldwide have Alzheimer’s Disease (AD), the most prevalent form of dementia [2], and this number is projected to reach over 150 million by 2050 [3]. Cognitive decline, even without dementia, increases the need for costly personal and medical care.

While extensive research has focused on risk factors for later-life cognitive decline and dementia, there are still remaining questions regarding its etiology. For example, AD is a result of the accumulation of amyloid beta plaques and neurofibrillary tangles in the brain [4]. Amyloid beta and tau protein metabolism may be influenced by genetic variants that alter chemical properties or abundance of relevant proteins [5]. Heritability estimates for AD are high (60-80%) [6], indicating that the identification of AD-associated variants is critical for a deeper etiological understanding. Heritability of cognitive function is also relatively high across the life course (40-80%) [7]. However, the vast majority of genetic loci for measures of cognitive function and dementia were identified from studies conducted in European Ancestry (EA) participants. A deeper exploration of the genetic factors underlying late-life cognition and dementia in non-EA populations is now needed to both identify population-specific risk variants across the genome and gauge the relative importance of previously-identified loci.

With over 1.4 billion people, India is the second most populous country in the world, and the public health burden of dementia is dramatically increasing as the population both grows and ages. The prevalence of dementia among South Asians living in India varies by geographic location and sociodemographic characteristics (e.g., rural vs. urban), and is approximately 7.4% among individuals 60 years and older [8]. While studies have indicated that older age, lower education, diabetes, obesity, and other factors increase risk of dementia in India [9], there has been little research on genetic risk factors. Therefore, it is unclear whether the same genes that have been associated with dementia and cognitive decline in EA have a similar influence on dementia risk in South Asians. Likewise, there may be causal risk variants in known AD genes, or in other genes, that are unique to India.

Detection of rare variants associated with measures of cognitive function in non-EA populations may be difficult due the combination of increased genetic diversity and smaller sample sizes available for genetic research, which leads to a loss of power. Statistical power can be increased by grouping together variants within a gene or genomic region that have the same functional annotation, such as those that alter protein structure (e.g. loss of function (LoF) or missense variants) or those in regulatory elements (e.g. gene promoter or enhancer regions), which helps increase the likelihood of selecting probable causal variants [10].

In this study, we examined whether 84 genes previously associated with AD in EA are also associated with seven measures of cognitive function in 2,680 participants from the Diagnostic Assessment of Dementia for Longitudinal Aging Study of India (LASI-DAD), a nationally-representative study that includes diverse ethno-linguistic and geographic groups. From whole genome sequence (WGS) data, we selected missense/loss-of-function (LoF) single-nucleotide variants (SNVs) and brain-specific promoter and enhancer SNVs within each gene. This work will help elucidate genetic variants associated with cognitive function in South Asians across India, which may play an important role in risk stratification and help guide intervention and treatment plans for those at risk for dementia in India.

## Methods

### Study population

LASI [11] is a nationally representative cohort of Indian adults who are at least 45 years of age. LASI-DAD, an ancillary study investigating risk factors for dementia, enrolled 4,096 LASI participants from 18 states and union territories across India. Participants were selected by two-stage stratified random sampling across states/territories in India and with respect to cognitive impairment risk, with sampling strategy described elsewhere [12,13]. Briefly, participants were classified as low risk or high risk for cognitive impairment based on their performance on core cognitive tests conducted in the larger LASI cohort, or on proxy reports if the participant did not complete those tests. Then, an approximately equal number of respondents in the high risk and low risk strata were randomly drawn from each state/territory with a target sample size proportionate to the population size. Participants underwent neurocognitive testing with tests logically and culturally adapted from tests present in the Harmonized Cognitive Assessment Protocol (HCAP) [12], informational interviews, and a blood draw to extract DNA for whole genome sequencing. A total of 2,680 participants with complete genotype and cognition data were included in the analysis.

### Whole-genome sequence data

Whole genome sequencing (WGS) at an average read depth of 30x was performed by MedGenome, Inc (Bangalore, India) using DNA extracted from blood samples from 2,762 LASI-DAD participants. Genotype calling and quality control (QC) were performed at the Genome Center for Alzheimer’s Disease (GCAD) at the University of Pennsylvania [14]. Briefly, sample-level quality control included checks for low coverage, sample contamination, sex discrepancies, concordance with previous genotype data, and duplicates [14]. After excluding control samples and samples with low quality and/or unresolved identity, a total of 2,680 samples were retained in the analysis. At the genotype level, each genotype was evaluated and set to missing if read depth was less than 10 (DP<10) or genotype quality score was less than 20 (GQ<20). At the variant level, a variant was excluded if it was monomorphic, was above the 99.8% Variant Quality Score Recalibration (VQSR) Tranche (the quality score was beyond the range that contains 99.8% of true variants), had a call rate ≤ 80%, or had an average mean depth > 500 reads. We further removed variants that were in low complexity regions identified with the mdust program.[15] After quality control and filtering, we retained a total of 71,109,961 autosomal bi-allelic variants that include 66,204,161 single nucleotide polymorphisms (SNPs) and 4,905,800 indels.

### Principal component analysis and genetic relationship matrix

We estimated genetic principal components (PCs) and the genetic relationship matrix (GRM) in GENESIS (version 2.26.0) [16,17]. For this analysis, we included variants with minor allele frequency (MAF) ≥ 5% and pruned for LD (r^2^=0.1, window size=500kb) to select independent variants. Kinship coefficients were first estimated using “snpgdsIBDKING” function. Subsequently, genetic principal components (PCs) were calculated using “PCair”, which estimates population structure while accounting for cryptic relatedness in the samples. Specifically, PCs were first estimated in a set of unrelated individuals (kinship cutoff = 0.044) to obtain robust variant weights, which were then used to project PCs in the rest of the sample. Following this, the genetic relationship matrix (GRM) was estimated using “PCrelate” by simultaneously adjusting the top 2 PCs to avoid potential confounding from population structure.

### Measures of Cognitive Function

We analyzed seven measures of cognitive function including five cognitive domains (memory, orientation, language/fluency, executive function, and visuospatial function), general cognitive function constructed from the five cognitive domain scores, and the Hindi Mental State Exam (HMSE) score. The HMSE is a version of the Mini Mental State Exam dementia screener translated into Hindi. It is designed to be administered to participants from a population where a significant proportion of individuals are illiterate, and is scored as the sum of 22 items which totals to an integer between zero and 30, with a higher score indicating more cognitive intactness [18]. The five cognitive domain scores are factor scores of a collection of tests assigned to a broad domain of cognition as informed by Cattell-Horn-Carroll (CHC) theory of human cognitive abilities, with composite weights and tests described elsewhere [19]. The cognitive domain and general cognitive function scores were each estimated using item-response theory (IRT) and were normalized to a Gaussian distribution with mean of zero and variance of one in the full LASI-DAD subcohort [19].

### Gene selection

We selected a total of 84 genes from the two largest genome-wide association studies (GWAS) for AD in EA (S1 Table) [20,21], as well as *TOMM40* and *APOC1* which are proximal to *APOE* and are known to be associated with Alzheimer’s disease [22,23]. Briefly, Bellenguez et al. [20] performed a two-stage GWAS of 10 case-control studies across Europe. Stage 1 included 39,106 clinically diagnosed AD cases, 46,828 proxy AD and dementia-related disorder (ADD) cases, and 401,577 controls. Stage 2 included 25,392 AD cases and 276,086 controls. Bellenguez et al. identified 75 risk loci, of which 42 were novel. The authors then conducted pathway analysis and designed a gene prioritization algorithm to stratify loci according to their likelihood of having a causal effect on ADD risk. For this study, we selected a total of 73 genes from Bellenguez et al. [20] including those that were labeled as known loci or those that were classified as having the highest likelihood of a causal effect on ADD (tier 1). Wightman et al. [21] conducted a meta-analysis of 13 studies of EA participants across the United States and Europe. The total sample size was 1,126,563 individuals, which included 90,338 AD cases (46,613 were proxy cases) and 1,036,225 controls (318,246 were proxy controls). For this study, we selected 45 genes from independent 38 loci that reached genome-wide significance (p<5×10^−8^) in Wightman et al. There was an overlap of 36 genes between the two AD GWAS. See Supplemental Methods for additional details on selecting genes within the identified AD loci (S1 Methods).

Gene boundaries were defined by GRCh38.p14 in NCBI Gene, which uses NCBI RefSeq to annotate gene positions. We selected all SNVs within the gene start and stop positions for the missense/LoF analysis. For the brain-specific promoter/enhancer analysis, we selected SNVs within a ± 20 kb buffer of the gene’s transcription start site. Only genes with at least two missense/LoF or promoter/enhancer SNVs (defined below) within the region were included in the final analysis.

### Definition of missense/LoF and promoter/enhancer SNVs

We followed similar definitions of missense/LoF SNVs, promoter SNVs, and enhancer SNVs as Li et al.[24] Briefly, we used the Variant Effect Predictor (VEP) [25] and LOFTEE [26] with GENCODE as the transcript annotation reference to identify missense and LoF SNVs, respectively. We additionally classified missense and LoF variants based on the confidence of their predicted function. LoF SNVs were annotated as either high confidence or low confidence using LOFTEE, and missense SNVs were assigned a REVEL score. REVEL scores are generated through an ensemble method to measure the pathogenicity of a missense SNV [27], with higher scores indicating greater likelihood of causing diseases. Missense SNVs with REVEL score>0.5 are considered to have high confidence. Next, we used the WGS Annotator (WGSA) v0.95 pipeline [28] to define promoter SNVs as those that fell within ± 5kb of a gene’s transcription start site with at least one H3K4me3 annotation for brain tissues (E067-E074, E081, E082) from the ENCODE database. We defined enhancer SNVs as those that fell within ± 20kb of the gene’s transcription start site and overlapped with an enhancer defined by EnhancerFinder in the brain.

### Annotation selection

For both the missense/LoF and promoter/enhancer analyses, we included all SNVs that had no missing annotation weights, regardless of minor allele frequency (S2 Methods). Annotation weights were retrieved using WGSA v0.95 [28]. We selected a subset of annotations similar to Li et al. [24], including those that predicted deleteriousness, predicted impact on the protein, and that summarized evolutionary conservation. For missense/LoF SNVs, we used CADD_raw_rankscore, a measure of variant deleteriousness combining multiple genomic features of each variant [29]; GERP_RS_rankscore, a measure of variant conservation [30]; Eigen.phred, a measure of variant deleteriousness using an unsupervised learning method [31]; and fathmm.MKL_coding_rankscore, a score from a machine learning method incorporating other annotations to predict deleteriousness of the variant from coding variants [32]. For promoter/enhancer SNVs, we used CADD_raw_rankscore [29], GERP_RS_rankscore [30], Eigen.PC.phred [31], fathmm.MKL_non.coding_rankscore [32], and GenoCanyon_rankscore, a measure of variant conservation [33]. Genome-wide ranks of the associated annotation scores were used to generate the Phred scores (i.e., the logarithmically transformed annotation score percentiles) required for STAAR analysis.

### Statistical methods

All analyses were conducted in R (ver. 4.2.0). WGS data were converted from VCF files to SeqArray GDS format using SNPRelate [34] and SeqArray [35] R packages. We then used the variant-Set Test for Association using Annotation infoRmation (STAAR) v0.9.6.1 to perform gene-based analysis using functional annotations for missense/LoF (including all variants except low-confidence LoF), missense/LoF (high confidence variants only), and promoter/enhancer regions separately [36]. In STAAR, linear mixed models were used to test each gene region for association with each of the seven measures of cognitive function separately, both with and without annotation weights. Model 1 adjusted for age, sex, state or union territory, and the first ten principal components of global ancestry. Model 2 additionally adjusted for educational attainment, rural or urban residence, and literacy status (yes/no). Each model incorporated a genetic relatedness matrix to account for relatedness between subjects, and geographic state or union territory was used to define heterogeneous variances within the linear mixed model.

For each gene, both with and without including annotation weights for the SNVs, we examined the STAAR p-value which is calculated from modified SKAT-(1,1), SKAT-(1,25), Burden-(1,1), Burden-(1,25), ACAT-V(1,1) and ACAT-V(1,25) tests. For analyses with and without annotation weights, separately, a Benjamini-Hochberg FDR q<0.1 was used to declare significance.

For each gene region that was associated with a measure of cognitive function at FDR q<0,1, we next performed a single variant analysis to identify the variants most strongly associated using a score test in STAAR. The same models from the gene-based analysis were used for single variant analysis. For the SNV with the lowest p-value within each identified gene, we compared the allele frequency in LASI-DAD to that found in EA samples registered in gnomAD v3 [37] to examine whether risk alleles were enriched in LASI-DAD.

## Results

The LASI-DAD analytic sample had a mean age of 69.6 (SD=7.3) years (Table 1). The majority of participants could not read or write (56.4%), lived in rural areas (63.3%), and had less than lower secondary education (75%). Mean HMSE score was 22.7 (SD=5.4) (S2 Table).

**Table 1:** Characteristics of the LASI-DAD analytic sample (N=2,680)

| Covariate | Count (%) or Mean (SD) |
| --- | --- |
| Age (years) | 69.6 (7.3) |
| Sex (female) | 1408 (52.5) |
| Literacy (cannot read or write) | 1511 (56.4) |
| Location |  |
| Rural | 1697 (63.3) |
| Urban | 983 (36.7) |
| Education |  |
| Less than lower secondary | 2004 (75) |
| Upper secondary & vocational training | 578 (22) |
| Tertiary | 98 (4) |
| HMSE score | 22.7 (5.4) |

HMSE = Hindi Mental State Exam, LASI-DAD = Longitudinal Aging Study in India – Diagnostic Assessment of Dementia

We next characterized the distribution of the annotation weights across missense/LoF SNVs and promoter/enhancer SNVs. The missense/LoF SNVs exhibited relatively little variation for almost all annotations and tended to be high. For these annotations, which were on a scale from 0 to 1, the median score ranged from 0.97 to 0.99 (S3 Table). The promoter/enhancer SNVs showed greater variation across annotation weights and tended to be lower (S4 Table). The Eigen-Phred and Eigen-PC-Phred rank scores had more variation and relatively low weights for missense variants, but low variation and higher weights for promoter/enhancer variants, due to their calculation with different training data for each functional class of variant (S Methods).

### Missense/loss-of-function (LoF) analysis

Of the 84 genes selected for analysis, 79 had at least two missense/LoF SNVs with complete annotations, and the median number of missense/LoF SNVs across the genes was 21 (Table 2). In Model 1, 16 genes were nominally associated with at least one measure of cognitive function (p<0.05, S5 Table), with 3 genes associated at FDR q<0.1 in the analysis without annotation weights (Table 3). Specifically, *APOE* was associated with HMSE score (FDR q=0.08), general cognitive function (FDR q=0.04), executive function (FDR q=0.07), and orientation (FDR q=0.07). *PICALM* was associated with HMSE score (FDR q=0.08), and *TSPOAP1* was associated with executive function (FDR q=0.07). In Model 2, which additionally adjusts for rural/urban location, literacy, and education, 20 genes were nominally associated with at least one measure of cognitive function (p<0.05, S6 Table), and *PICALM* was significantly associated with HMSE score after correction for multiple testing (FDR q=0.098) in the analysis without annotation weights (Table 3).

**Table 2:** Five-number summary of variants in Missense/Loss-of-Function and Promoter/Enhancer analyses.

| Analysis | Min. | Q1 | Median | Q3 | Max. | Number of Genes | # SNVs With MAF > 0 | Total # SNVs Analyzed* |
| --- | --- | --- | --- | --- | --- | --- | --- | --- |
| Missense/LoF | 2 | 14.5 | 22 | 39 | 178 | 79 | 2515 | 2512 |
| Missense | 2 | 14 | 21 | 38 | 167 | 79 | 2447 | 2444 |
| LoF | 1 | 1 | 1 | 2 | 11 | 36 | 68 | 68 |
| Promoter/Enhancer | 6 | 61 | 93 | 127 | 265 | 77 | 7402 | 7370 |
| Promoter | 6 | 59 | 91 | 125 | 231 | 77 | 7108 | 7077 |
| Enhancer | 29 | 37.25 | 48 | 62 | 88 | 10 | 509 | 508 |

SNV = single nucleotide variant, MAF = minor allele frequency, LoF = loss-of-function, Q1 = quartile 1, Q3 = quartile 3.
\*SNVs were analyzed if they met the definition for inclusion for the analysis indicated, had a MAF > 0, and had a complete set of annotation weights.
Total number of SNVs is the number of unique SNVs analyzed across all genes selected for analysis.

**Table 3:**
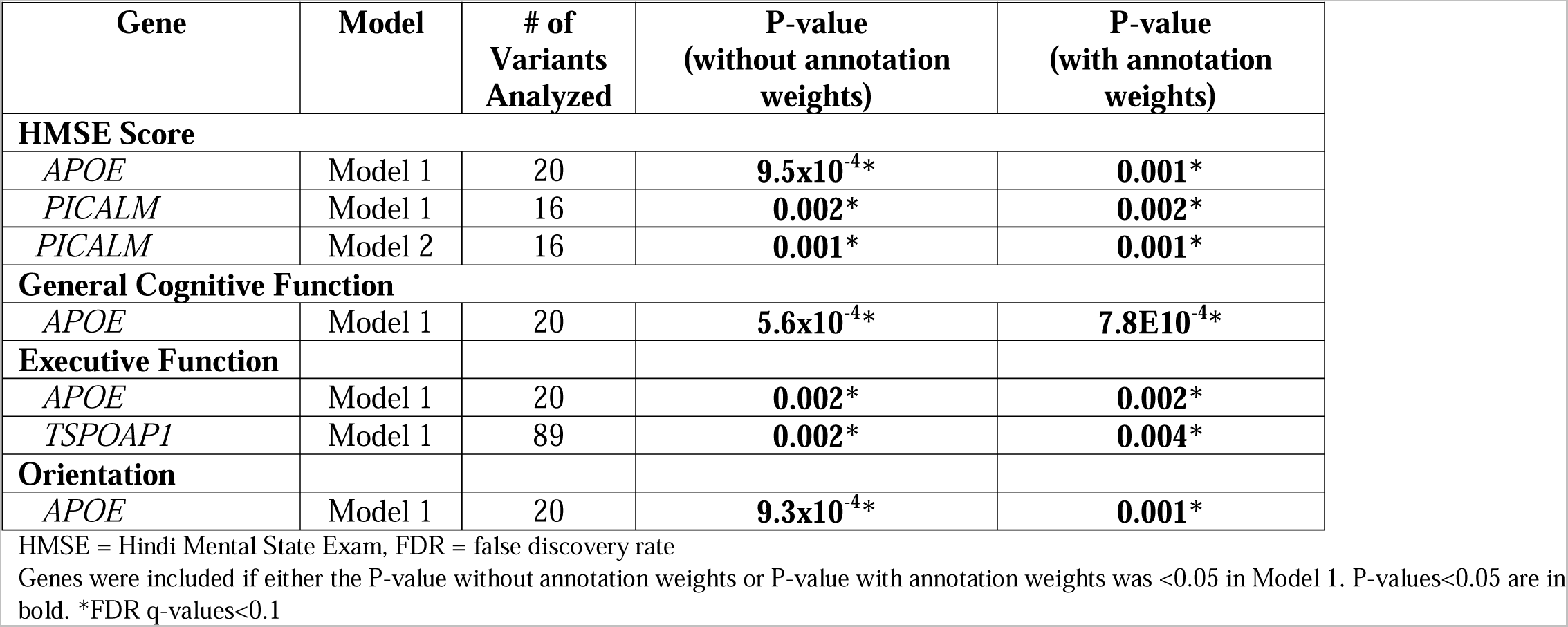
Genes with FDR q<0.1 in Missense/LoF Analysis.

As shown in Table 3, the results were similar when we used annotation weights. At FDR q<0.1, *APOE* was associated with HMSE score (FDR q=0.08) and general cognitive function (FDR q=0.07), and *PICALM* was associated with HMSE score (FDR q=0.08). In Model 2, at FDR q<0.1, *PICALM* was associated with HMSE score (FDR q=0.09).

For each gene associated with a cognitive measure at FDR q<0.1, we examined associations between each SNV within the gene region, without annotation weights, and the cognitive outcome of interest (Table 4). As expected, the most strongly associated variant in Model 1 within *APOE* for all measures of cognitive function was rs429358 (HMSE p=2.9×10^−4^, general cognitive function p=1.4×10^−4^, executive function p=4.1×10^−4^, orientation p=2.4×10^−4^; Fig 1, S1-S3 Figs), which is the missense variant in exon 4 of *APOE* that changes cysteine to arginine and differentiates the *APOE* ε4 allele from ε2 and ε3. Removal of this SNV results in *APOE* losing significance. For *TSPOAP1* in Model 1, the most strongly associated variant with executive function was rs9913145 (Model 1 p=5.7×10^−4^), a missense variant in exon 17 that changes glutamine to arginine (Fig 2). This variant had a MAF of 0.15 in LASI-DAD, and a MAF of 0.12 in EA samples in gnomAD, which indicates that the minor allele is relatively common both in LASI-DAD and in EA populations. In *PICALM* in Model 1 and Model 2, the most strongly associated SNV with HMSE was rs779406084 (Model 1 p=4.2×10^−4^, Model 2 p=1.6×10^−4^), a missense variant in exon 19 that changes threonine to methionine (Fig 3, S4 Fig). Rs779406084 was in high LD with all missense/LoF SNVs in *PICALM* (|D’|=1) but was not correlated with any other missense/LoF SNV in the gene (r^2^<<0.2) including with the one other missense/LoF SNV with p<0.05. This SNV has a CADD score of 24, indicating that it is in the top 0.4^th^ percentile of all deleterious SNVs. It also has a MAF of 7.5×10^−4^ in LASI-DAD. While very rare, this variant occurs more often in LASI-DAD compared to EA samples in gnomAD (EA MAF=1.5×10^−5^).

**Figure 1.**
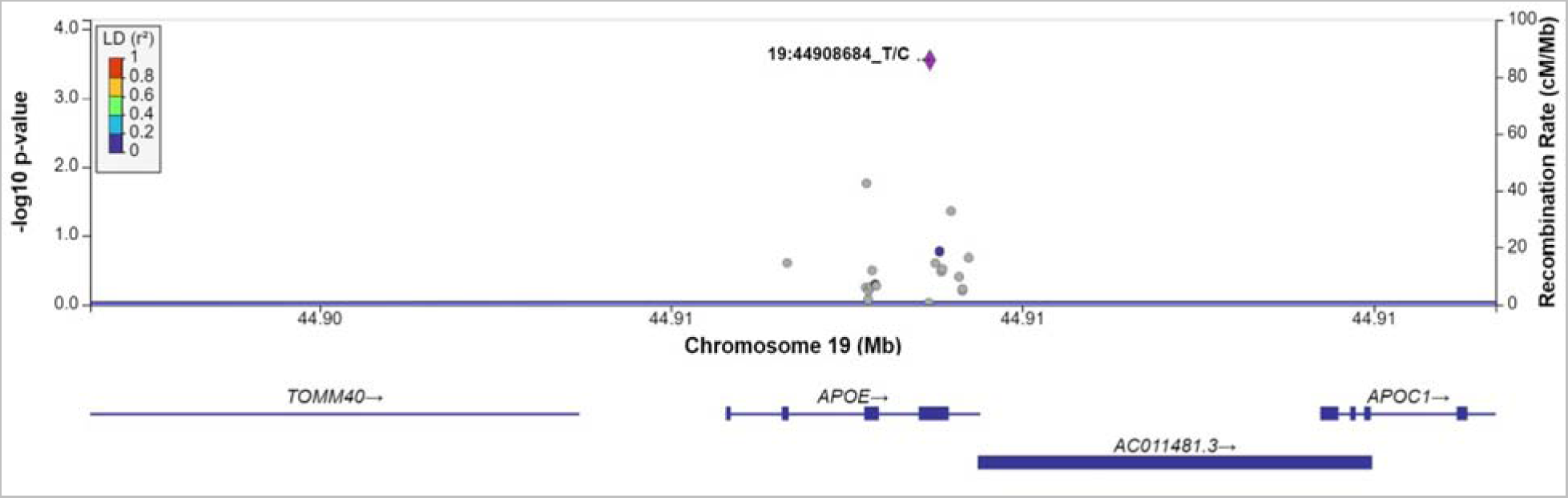
Plot of missense/loss-of-function SNVs in *APOE* in Model 1 for Hindi Mental State Exam (HMSE) score with no annotation weights. Left Y-axis: −log10(p-value) from association between SNV and HMSE score, adjusting for age, sex, state/territory, the first 10 principal components of genetic ancestry, and accounting for relatedness (random effect) and heteroscedastic variances among state/territory; Right Y-axis: SNV recombination rate based on HapMap GRCh38 South Asian sample (SAS); X-axis: chromosomal location and gene regions; LD r^2^ color code: degree of linkage disequilibrium with index (most strongly associated) SNV, rs429358 (purple diamond). Grey points indicate no LD information present in the reference panel.

**Figure 2.**
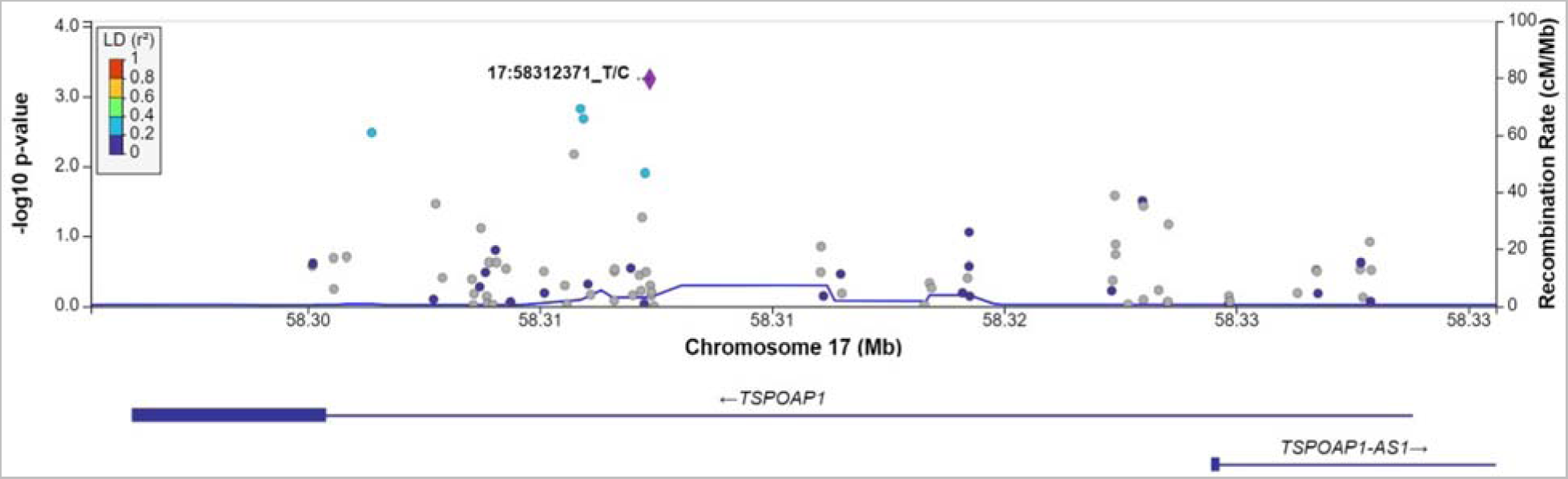
Plot of missense/loss-of-function SNVs in *TSPOAP1* gene in Model 1 for executive function. Left Y-axis: −log10(p-value) from association between SNV and executive function, adjusting for age, sex, state/territory, the first 10 principal components of genetic ancestry; and accounting for relatedness and heteroscedastic variances among state/territory; Right Y-axis: SNV recombination rate based on HapMap GRCh38 SAS; X-axis: chromosomal location and gene regions; r^2^ color code: degree of linkage disequilibrium with index (most strongly associated) SNV, rs9913145 (purple diamond). Grey points indicate no LD information present in the reference panel. No annotation weights were used to generate p-value.

**Figure 3.**
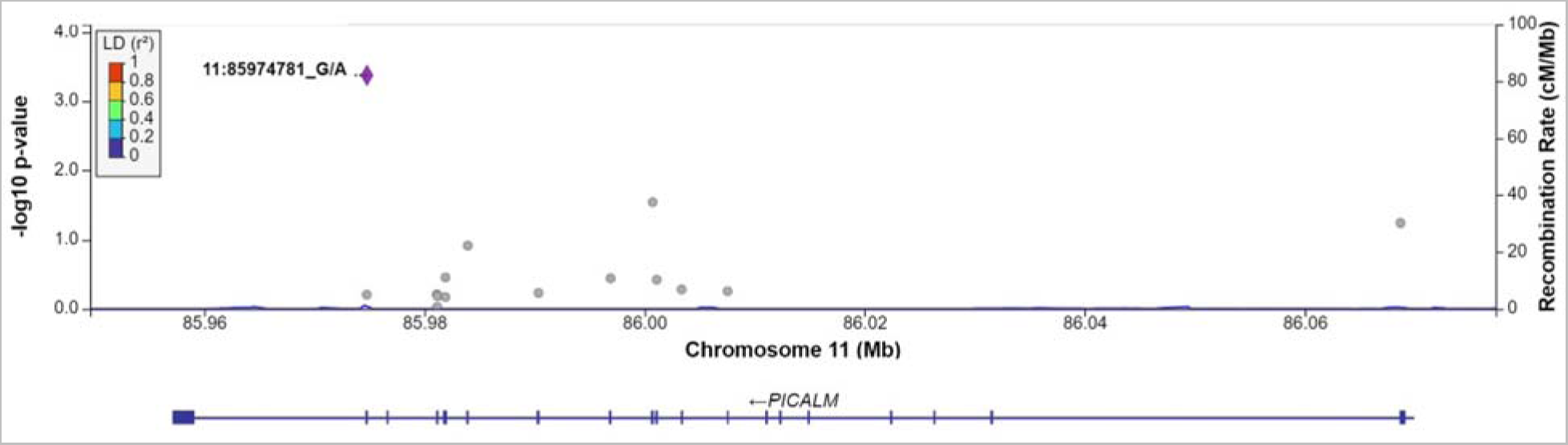
Plot of missense/loss-of-function SNVs in *PICALM* gene in Model 1 for HMSE score. Left Y-axis: −log10(p-value) from association between SNV and Hindi Mental State Exam (HMSE) score, adjusting for age, sex, state/territory, the first 10 principal components of genetic ancestry; and accounting for relatedness and heteroscedastic variances among state/territory; Right Y-axis: SNV recombination rate based on HapMap GRCh38 SAS; X-axis: chromosomal location and gene regions; r^2^ color code: degree of linkage disequilibrium with index (most strongly associated) SNV, rs779406084 (purple diamond). Grey points indicate no LD information present in the reference panel. No annotation weights were used to generate p-values.

**Table 4:**
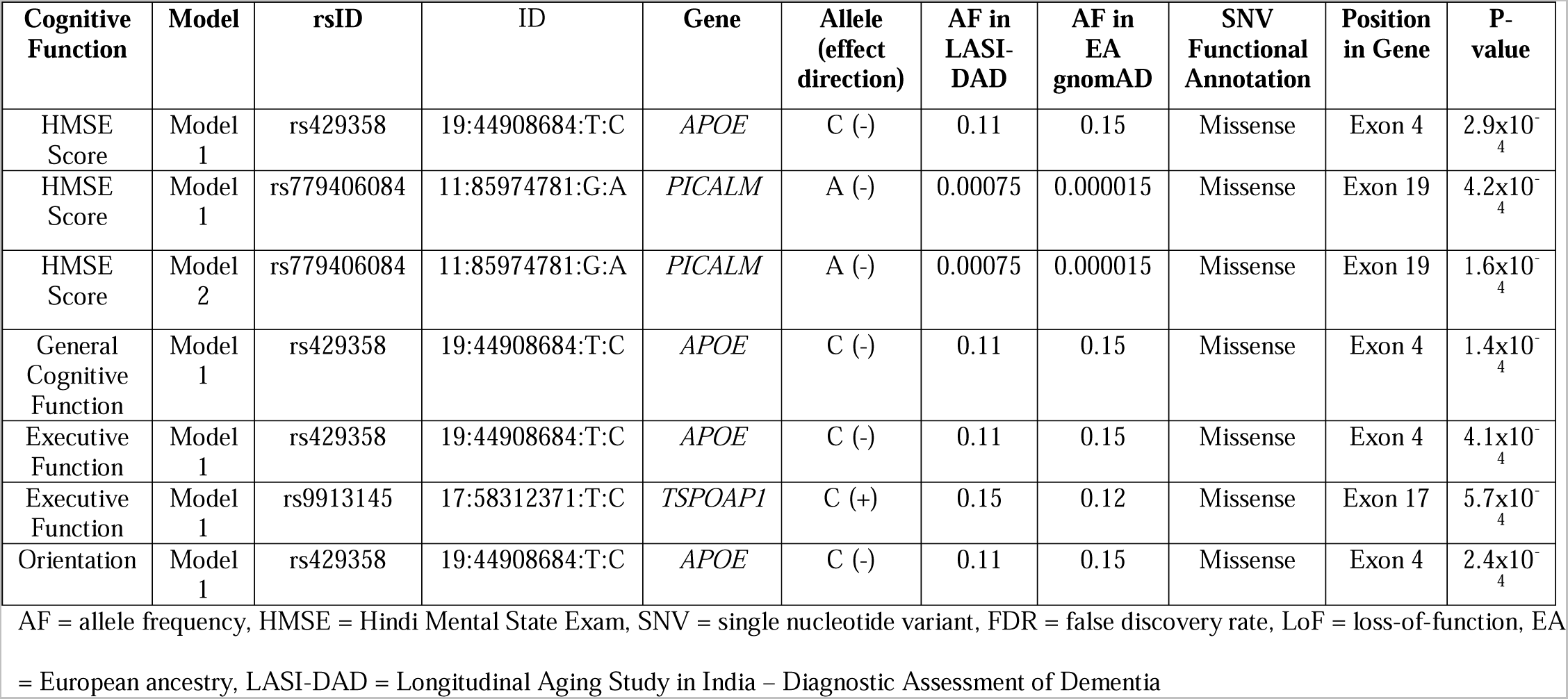
Sentinel SNVs from genes with FDR q<0.1 in Model 1 or Model 2 of the Missense/LoF analysis without annotation weights.

### High-confidence missense/LoF analysis

Of the 84 genes analyzed, 51 had at least two high-confidence LoF or missense SNVs with REVEL>0.5. In Model 1, 8 genes were nominally associated with at least one measure of cognitive function (p<0.05, S7 Table), but none were significant after correction for multiple testing (FDR q>0.1). In Model 2, 10 genes were also nominally associated with at least one measure of cognitive function (p<0.05, S8 Table) with four genes overlapping from Model 1 (*ABI3, APOE*, *DGKQ,* and *INPP5D*), but none were significant after correction for multiple testing (FDR q>0.1). For both analyses, the results did not change substantively when annotation weights were incorporated.

### Promoter/enhancer analysis

Of the 84 genes analyzed, 77 had at least two brain-specific promoter or enhancer SNVs with complete annotation weights, and the median number of brain-specific promoter/enhancer SNVs across the genes was 93 (Table 2). In Models 1 and 2, 21 and 22 genes, respectively, were nominally associated with at least one measure of cognitive function without annotation weights (p<0.05), but none were associated after multiple testing correction (all FDR q>0.1, S9 and S10 Tables). When we incorporated annotation weights, 18 genes in Model 1 and 22 genes in Model 2 were nominally associated with at least one measure of cognitive function, but again none were associated after multiple testing correction (all FDR q>0.1, S9 and S10 Tables), with 18 and 20 genes being nominally associated with at least one measure of cognitive function with and without annotation weights, respectively. In Model 1, *USP6NL*, *INPP5D*, and *KAT8* were no longer nominally associated with any measure of cognitive function after incorporation of annotation weights. In Model 2, *EGFR* and *APOE* were no longer nominally associated with any measure of cognitive function after incorporation of annotation weights, but *APOC1* and *SORT1* became nominally associated.

## Discussion

We performed a gene-based analysis examining the association between missense/LoF and brain-specific enhancer and promoter variants in previously-identified AD-associated genes and seven measures of cognitive function in South Asians across India. Using only missense/LoF variants, three genes were associated with at least one measure of cognitive function after multiple testing correction, including *APOE* with multiple cognitive measures. However, no genes were associated in the brain-specific promoter and enhancer analysis. The most strongly associated variants were missense SNVs with high predicted deleteriousness. One of the most significantly associated missense variants was very rare, yet appeared to be enriched in LASI-DAD compared to EA samples in public databases.

We found that *APOE* is significantly associated with HMSE score, general cognitive function, executive function, and orientation in the missense/LoF analysis in Model 1. Apolipoprotein E (*APOE)* facilitates cholesterol and phospholipid transfer between cells, and complexes with amyloid β proteins in the brain for removal, inhibiting the amyloid β plaque formation necessary for AD onset [38]. *APOE* alleles confer different risks for Alzheimer’s disease. Relative to the ε3 allele, ε4 is associated with increased risk of Alzheimer’s disease in EA [39], and the ε4/ε4 genotype is also associated with cognitive decline in those with Alzheimer’s disease [40]. In our analytic sample from LASI-DAD, the ε4 allele frequency is estimated to be approximately 10.9%, which is less than reported frequency among EA samples in the US (14%) [41] and while not common is still frequent. The ε4 allele has somewhat different associations with AD risk across races/ethnicities, with ε4 and associated variant effects being stronger in EA populations compared to African-Americans [42]. Rs429358 is used to differentiate between ε3 and ε4 alleles in *APOE*, and has a CADD score of 16.6, which places it in the top 2^nd^ percentile of deleterious SNVs. Although previous studies with a subset of the current LASI-DAD sample did not find an association between cognitive function and rs429358 [17,43], this was likely due to smaller sample size and/or less regional variation in the previous studies. Our reported associations with *APOE* are not surprising as working memory and executive function deficits are often early markers of AD [44].

Phosphatidylinositol binding clathrin assembly protein (*PICALM*) facilitates endocytosis of APP [45], which is needed to form β-amyloid plaques that lead to AD. *PICALM* was found to be associated with cognitive function in EA samples [46]. *PICALM* variants identified in EA have had mixed associations in East Asian samples, with which the South Asian population of India shares ancestry [47]. For example, some variants identified as associated with AD in a large meta-analysis of Chinese GWAS near *PICALM* [48]were not found to be associated in smaller Indian studies [49]. We found that the sentinel SNV in *PICALM* for HMSE score is rs779406084, a very rare variant with a high CADD score at 24, placing it in the top 0.2^th^ percentile of all deleterious SNVs. To our knowledge, this is the first study that reports an association with rs779406084 and cognitive function. This is likely due to the rarity of this variant in EA samples (MAF=1.5×10^−5^) and an association was likely found in our study due to its comparatively higher frequency in LASI-DAD (MAF=7.5×10^−4^). Further work is needed to elucidate the specific effects of this variant on the protein and replication in other cohorts is needed.

Translocator protein (TSPO) associated protein 1 (*TSPOAP1*) regulates calcium channels in nerve synapses [50]. It interacts with the protein TSPO which is involved in inflammation pathways [51]. *TSPOAP1* variants were associated with AD in a large transethnic AD GWAS [52]. The sentinel SNV of *TSPOAP1* in LASI-DAD was rs9913145, which has a CADD score of 1.12 indicating that it is not strongly deleterious. This variant is slightly more common in LASI-DAD (MAF=0.15) compared to EA samples (MAF=0.12). To our knowledge, this variant has not otherwise been reported to have an association with cognitive function or dementia. Given the relatively low CADD score of the variant and the relatively common frequency in EA samples, this variant may tag a haplotype specific to South Asians within *TSPOAP1* that is associated with executive function.

We found no associations between brain-specific promoter and enhancer SNVs within the known AD genes and any of the measures of cognitive function in our sample after multiple testing correction. This is likely due to promoter/enhancer SNVs having more subtle effects on AD gene expression compared to the potentially more deleterious effects from missense/LoF SNVs. We also found that annotation weights did not substantively change our analysis results. This may be because the missense/LoF variants had relatively small variance in their annotation weights and tended to be high, resulting in little additional statistical information.

We found many genes that were nominally associated with each measure of cognitive function in the missense/LoF analysis and brain-specific promoter/enhancer analysis. Genes associated with multiple cognitive measures include *ADAM17, OTULIN,* and *ABCA7,* which are involved in amyloid-β metabolism or in immune signaling [38,53,54]. In both Model 1 and Model 2, *ABCA7, ADAM17, APOE, OTULIN,* and *TSPOAP1* were all at least nominally associated with three or more measures of cognitive function. These genes all play a role in cholesterol and APP metabolism (*ABCA7, ADAM17, APOE)* or are involved in inflammation pathways (*OTULIN, TSPOAP1*).

Similar gene-based analysis studies were conducted in EA samples. One large genome-wide gene-based AD study conducted in the UK BioBank on different categories of rare missense/LoF variants found that three gene regions were associated with AD parent proxy cases, including *TOMM40/APOE* [55]. Notably, detection of these regions depended on resolving variants into categories of high confidence and predicted loss-of-function effects. Another gene-based AD analysis conducted in the ADES-FR study found that protein-truncating rare variants and strictly damaging rare variants in *TREM2*, *ABCA7*, and *SORL1* were associated with early-onset AD, but not with late onset AD [56]. Given that the previous studies focused on very specific classes of rare variants in EA samples, it is no surprise that these genes were also at least nominally associated in our study.

Although many genes were nominally associated in our analysis, few genes were significant after correction for multiple testing. This could be in part because we selected genes associated with AD, which may have weaker effects on cognitive function changes that precede AD. Further, the genes were identified through GWAS which excels in identifying primarily non-functional common variants which may be correlated with causal variants. In this study, >95% of our analyzed variants were rare (MAF<5%). Although we focused on variants more likely to be causal, it is possible that the sentinel SNPs identified in the GWAS were not tagging variants in the functional classes we examined, and that more genes would have reached significance if common, non-functional variation was included in our analysis. Further, the genes identified were in large cohorts of EA. Genetic differences between EA and South Asians, including allele frequency and linkage disequilibrium, could have contributed to the lack of findings. Another explanation is that genetic associations with cognitive function may be attenuated in this population due to more heterogeneity in environmental factors across India, such as sociodemographics, socio-cultural factors, and air pollution, each of which is associated with cognitive function [57–61]. Finally, the tendency toward associations being nominally significant, but not significant after multiple testing correction, may be a result of the smaller sample size.

One limitation of this study is that we examined only two classes of functional annotations. It may be that variants with other functional consequences besides missense/LoF and promoter/enhancer variants could influence associations with cognitive function. Another limitation is that we could not include insertion/deletion variants in this analysis, as annotation weights are not available; however, it is possible that these variants may have more deleterious effects on proteins and their removal may have attenuated signal. Additionally, the LASI-DAD cohort design oversamples LASI participants with higher cognitive impairment risk, which may result in different observed genetic associations with cognitive function compared to studies sampled in other ways [17]. Finally, cognitive measures may have been biased due to administering the tests in many different languages [43]. However, no systematic bias with respect to language has been detected in LASI-DAD [62].

Our study also has several strengths. The prioritization and aggregation of SNVs based on their actual or predicted functional consequences likely increased signal for associations between the genes and cognitive function by focusing on variants that are more likely to have causal effects. Gene-based analysis with functional annotation also more directly links SNVs disease etiology, allowing a greater understanding of the types of variation within these genes that contribute to cognitive function. Additionally, to our knowledge, our study is the first to examine gene-based, rare variant associations with cognitive function in South Asians living in India. Thus, this work addresses an important health disparity in an understudied population [63]. Furthermore, our cohort presented a unique genetic environment to study potentially novel associations with understudied genetic variants due to its large genetic heterogeneity, unique subpopulations, and unique genetic ancestry [43,47,64,65]. Finally, we examined several measures of cognitive function, which allows us to determine which specific cognitive domains are associated with each gene.

In conclusion, we found that three genes (*APOE, PICALM,* and *TSPOAP1*) associated with Alzheimer’s disease in EA are also associated with measure of cognitive functions in South Asians living in India, with the association primarily driven by missense/LoF SNVs. Associations were in part driven by rare, deleterious alleles, including a very rare SNV enriched in LASI-DAD compared to EA. Future functional studies are needed to verify and characterize SNVs found within this study.

## Financial disclosure statement

This project is funded by the National Institute on Aging (R01 AG051125, U01 AG064948). The study sponsor had no role in the design and conduct of the study; in the collection, analysis, and interpretation of the data; or in the preparation, review, or approval of the manuscript.

## Competing interests

The authors have no conflicts of interest to disclose.

## Ethics statement

Informed consent was obtained from all participants, and the study was approved by Institutional Review Boards at the University of Southern California and the University of Michigan.

## Data availability

Genomic data can be accessed through The National Institute on Aging Genetics of Alzheimer’s Disease Data Storage Site (NIAGADS, accession number: NG00067.v10).

## Data Availability

Whole genome sequencing data for the Diagnostic Assessment of Dementia for the Longitudinal Aging Study of India (LASI-DAD) is available from the National Institute on Aging Genetics of Alzheimer's Disease Data Storage Site (NIAGADS), accession number: NG00067-ADSP Umbrella. Phenotype data is available at the Gateway to Global Aging website, https://g2aging.org/.

https://dss.niagads.org/datasets/ng00067/

https://g2aging.org/.

## Notes

### Competing Interest Statement

The authors have declared no competing interest.

## References

1. Wimo A, Seeher K, Cataldi R, Cyhlarova E, Dielemann JL, Frisell O, et al. The worldwide costs of dementia in 2019. Alzheimer’s Dement. 2023;n/a. 10.1002/alz.12901

2. Schneider JA, Arvanitakis Z, Leurgans SE, Bennett DA. The neuropathology of probable Alzheimer disease and mild cognitive impairment. Ann Neurol. 2009;66: 200–208. doi:10.1002/ana.21706

3. Breijyeh Z, Karaman R. Comprehensive Review on Alzheimer’s Disease: Causes and Treatment. Molecules. 2020;25. doi:10.3390/molecules25245789

4. Huber CM, Yee C, May T, Dhanala A, Mitchell CS. Cognitive Decline in Preclinical Alzheimer’s Disease: Amyloid-Beta versus Tauopathy. J Alzheimer’s Dis. 2018;61: 265–281. doi:10.3233/JAD-170490

5. Area-Gomez E, Schon EA. Alzheimer disease. Adv Exp Med Biol. 2017;997: 149–156. doi:10.1007/978-981-10-4567-7_11

6. Sims R, Hill M, Williams J. The multiplex model of the genetics of Alzheimer’s disease. Nat Neurosci. 2020;23: 311–322. doi:10.1038/s41593-020-0599-5

7. Mollon J, Knowles EEM, Mathias SR, Gur R, Peralta JM, Weiner DJ, et al. Genetic influence on cognitive development between childhood and adulthood. Mol Psychiatry. 2021;26: 656–665. doi:10.1038/s41380-018-0277-0

8. Lee J, Meijer E, Langa KM, Ganguli M, Varghese M, Banerjee J, et al. Prevalence of dementia in India: National and state estimates from a nationwide study. Alzheimer’s Dement. 2023;19: 2898–2912. doi:10.1002/alz.12928

9. Ravindranath V, Sundarakumar JS. Changing demography and the challenge of dementia in India. Nat Rev Neurol. 2021;17: 747–758. doi:10.1038/s41582-021-00565-x

10. Zuk O, Schaffner SF, Samocha K, Do R, Hechter E, Kathiresan S, et al. Searching for missing heritability: Designing rare variant association studies. Proc Natl Acad Sci U S A. 2014;111. doi:10.1073/pnas.1322563111

11. Perianayagam A, Bloom D, Lee J, Parasuraman S, Sekher T V, Mohanty SK, et al. Cohort Profile: The Longitudinal Ageing Study in India (LASI). Int J Epidemiol. 2022;51: e167–e176. doi:10.1093/ije/dyab266

12. Lee J, Khobragade PY, Banerjee J, Chien S, Angrisani M, Perianayagam A, et al. Design and Methodology of the Longitudinal Aging Study in India-Diagnostic Assessment of Dementia (LASI-DAD). J Am Geriatr Soc. 2020;68 Suppl 3: S5–S10. doi:10.1111/jgs.16737

13. Lee J, Dey AB. Introduction to LASI-DAD: The Longitudinal Aging Study in India-Diagnostic Assessment of Dementia. Journal of the American Geriatrics Society. United States; 2020. pp. S3–S4. doi:10.1111/jgs.16740

14. Leung YY, Valladares O, Chou YF, Lin HJ, Kuzma AB, Cantwell L, et al. VCPA: Genomic variant calling pipeline and data management tool for Alzheimer’s Disease Sequencing Project. Bioinformatics. 2019;35: 1768–1770. doi:10.1093/bioinformatics/bty894

15. Morgulis A, Gertz EM, Schäffer AA, Agarwala R. A Fast and Symmetric DUST Implementation to Mask Low-Complexity DNA Sequences. J Comput Biol. 2006;13: 1028–1040. doi:10.1089/cmb.2006.13.1028

16. Gogarten SM, Sofer T, Chen H, Yu C, Brody JA, Thornton TA, et al. Genetic association testing using the GENESIS R/Bioconductor package. Bioinformatics. 2019. doi:10.1093/bioinformatics/btz567

17. Smith JA, Zhao W, Yu M, Rumfelt KE, Moorjani P, Ganna A, et al. Association Between Episodic Memory and Genetic Risk Factors for Alzheimer’s Disease in South Asians from the Longitudinal Aging Study in India-Diagnostic Assessment of Dementia (LASI-DAD). J Am Geriatr Soc. 2020;68 Suppl 3: S45–S53. doi:10.1111/jgs.16735

18. Tsolaki M, Iakovidou V, Navrozidou H, Aminta M, Pantazi T, Kazis A. Hindi Mental State Examination (HMSE) as a screening test for illiterate demented patients. Int J Geriatr Psychiatry. 2000;15: 662–664. 10.1002/1099-1166(200007)15:7<662::AID-GPS171>3.0.CO;2-5

19. Gross A, Khobragade P, Meijer E, Saxton J. Measurement and Structure of Cognition in the Longitudinal Aging Study in India—Diagnostic Assessment of Dementia. Innovation in Aging. 2020. p. 660. doi:10.1093/geroni/igaa057.2280

20. Bellenguez C, Küçükali F, Jansen IE, Kleineidam L, Moreno-Grau S, Amin N, et al. New insights into the genetic etiology of Alzheimer’s disease and related dementias. Nat Genet. 2022;54: 412–436. doi:10.1038/s41588-022-01024-z

21. Wightman DP, Jansen IE, Savage JE, Shadrin AA, Bahrami S, Holland D, et al. A genome-wide association study with 1,126,563 individuals identifies new risk loci for Alzheimer’s disease. Nat Genet. 2021;53: 1276–1282. doi:10.1038/s41588-021-00921-z

22. Zhou Q, Zhao F, Lv ZP, Zheng CG, Zheng WD, Sun L, et al. Association between APOC1 polymorphism and alzheimer’s disease: A case-control study and meta-analysis. PLoS One. 2014;9. doi:10.1371/journal.pone.0087017

23. Corder EH, Saunders AM, Strittmatter WJ, Schmechel DE, Gaskell PC, Small GW, et al. Gene dose of apolipoprotein E type 4 allele and the risk of Alzheimer’s disease in late onset families. Science (80-). 1993;261: 921–923. doi:10.1126/science.8346443

24. Lee S, Shi B, Peloso GM, Wang Y, Heard-Costa N, Lin H, et al. Functional Annotations-Informed Whole Genome Sequence Analysis Identifies Novel Rare Variants for AD in the Alzheimer’s Disease Sequencing Project. Alzheimer’s Dement. 2022;18: e063968. 10.1002/alz.063968

25. McLaren W, Gil L, Hunt SE, Riat HS, Ritchie GRS, Thormann A, et al. The Ensembl Variant Effect Predictor. Genome Biol. 2016;17: 122. doi:10.1186/s13059-016-0974-4

26. Karczewski KJ, Francioli LC, Tiao G, Cummings BB, Alföldi J, Wang Q, et al. The mutational constraint spectrum quantified from variation in 141,456 humans. Nature. 2020;581: 434–443. doi:10.1038/s41586-020-2308-7

27. Ioannidis NM, Rothstein JH, Pejaver V, Middha S, McDonnell SK, Baheti S, et al. REVEL: An Ensemble Method for Predicting the Pathogenicity of Rare Missense Variants. Am J Hum Genet. 2016;99: 877–885. doi:10.1016/j.ajhg.2016.08.016

28. Liu X, White S, Peng B, Johnson AD, Brody JA, Li AH, et al. WGSA: An annotation pipeline for human genome sequencing studies. J Med Genet. 2015;53: 111–112. doi:10.1136/jmedgenet-2015-103423

29. Rentzsch P, Witten D, Cooper GM, Shendure J, Kircher M. CADD: Predicting the deleteriousness of variants throughout the human genome. Nucleic Acids Res. 2019;47: D886–D894. doi:10.1093/nar/gky1016

30. Davydov E V., Goode DL, Sirota M, Cooper GM, Sidow A, Batzoglou S. Identifying a high fraction of the human genome to be under selective constraint using GERP++. PLoS Comput Biol. 2010;6. doi:10.1371/journal.pcbi.1001025

31. Ionita-Laza I, Mccallum K, Buxbaum J. A SPECTRAL APPROACH INTEGRATING FUNCTIONAL GENOMIC ANNOTATIONS FOR CODING AND NONCODING VARIANTS IULIANA IONITA-LAZA HHS Public Access Author manuscript. Nat Genet. 2016;48: 214–220. doi:10.1038/ng.3477.A

32. Shihab HA, Rogers MF, Gough J, Mort M, Cooper DN, Day INM, et al. An integrative approach to predicting the functional effects of non-coding and coding sequence variation. Bioinformatics. 2015;31: 1536–1543. doi:10.1093/bioinformatics/btv009

33. Lu Q, Hu Y, Sun J, Cheng Y, Cheung KH, Zhao H. A statistical framework to predict functional non-coding regions in the human genome through integrated analysis of annotation data. Sci Rep. 2015;5: 1–13. doi:10.1038/srep10576

34. Zheng X, Levine D, Shen J, Gogarten S, Laurie C, Weir B. A High-performance Computing Toolset for Relatedness and Principal Component Analysis of SNP Data. Bioinformatics. 2012;28: 3326–3328. doi:10.1093/bioinformatics/bts606

35. Zheng X, Gogarten S, Lawrence M, Stilp A, Conomos M, Weir B, et al. SeqArray -- A storage-efficient high-performance data format for WGS variant calls. Bioinformatics. 2017. doi:10.1093/bioinformatics/btx145

36. Li X, Li Z, Zhou H, Gaynor SM, Liu Y, Chen H, et al. Dynamic incorporation of multiple in silico functional annotations empowers rare variant association analysis of large whole-genome sequencing studies at scale. Nat Genet. 2020;52: 969–983. doi:10.1038/s41588-020-0676-4

37. Chen S, Francioli LC, Goodrich JK, Collins RL, Kanai M, Wang Q, et al. A genome-wide mutational constraint map quantified from variation in 76,156 human genomes. bioRxiv. 2022; 2022.03.20.485034. doi:10.1101/2022.03.20.485034

38. Knopman DS, Amieva H, Petersen RC, Chételat G, Holtzman DM, Hyman BT, et al. Alzheimer disease. Nat Rev Dis Prim. 2021;7: 33. doi:10.1038/s41572-021-00269-y

39. scholar.

40. Martins CAR, Oulhaj A, de Jager CA, Williams JH. APOE alleles predict the rate of cognitive decline in Alzheimer disease. Neurology. 2005;65: 1888 LP–1893. doi:10.1212/01.wnl.0000188871.74093.12

41. Farrer LA, Cupples LA, Haines JL, Hyman B, Kukull WA, Mayeux R, et al. Effects of Age, Sex, and Ethnicity on the Association Between Apolipoprotein E Genotype and Alzheimer Disease: A Meta-analysis. JAMA. 1997;278: 1349–1356. doi:10.1001/jama.1997.03550160069041

42. Kulminski AM, Shu L, Loika Y, Nazarian A, Arbeev K, Ukraintseva S, et al. APOE region molecular signatures of Alzheimer’s disease across races/ethnicities. Neurobiol Aging. 2020;87: 141.e1–141.e8. doi:10.1016/j.neurobiolaging.2019.11.007

43. Zhao W, Smith JA, Wang YZ, Chintalapati M, Ammous F, Yu M, et al. Polygenic Risk Scores for Alzheimer’s Disease and General Cognitive Function Are Associated With Measures of Cognition in Older South Asians. Journals Gerontol Ser A. 2023;78: 743–752. doi:10.1093/gerona/glad057

44. Kirova A-M, Bays RB, Lagalwar S. Working memory and executive function decline across normal aging, mild cognitive impairment, and Alzheimer’s disease. Biomed Res Int. 2015;2015: 748212. doi:10.1155/2015/748212

45. Xu W, Tan L, Yu J-T. The Role of PICALM in Alzheimer’s Disease. Mol Neurobiol. 2015;52: 399–413. doi:10.1007/s12035-014-8878-3

46. Mengel-From J, Christensen K, McGue M, Christiansen L. Genetic variations in the CLU and PICALM genes are associated with cognitive function in the oldest old. Neurobiol Aging. 2011;32: 554.e7–554.e11. 10.1016/j.neurobiolaging.2010.07.016

47. Reich D, Thangaraj K, Patterson N, Price AL, Singh L. Reconstructing Indian population history. Nature. 2009;461: 489–494. doi:10.1038/nature08365

48. Liu G, Xu Y, Jiang Y, Zhang L, Feng R, Jiang Q. PICALM rs3851179 Variant Confers Susceptibility to Alzheimer’s Disease in Chinese Population. Mol Neurobiol. 2017;54: 3131–3136. doi:10.1007/s12035-016-9886-2

49. Shankarappa BM, Kota LN, Purushottam M, Nagpal K, Mukherjee O, Viswanath B, et al. Effect of CLU and PICALM polymorphisms on AD risk: A study from south India. Asian J Psychiatr. 2017;27: 7–11. doi:10.1016/j.ajp.2016.12.017

50. Mencacci NE, Brockmann MM, Dai J, Pajusalu S, Atasu B, Campos J, et al. Biallelic variants in TSPOAP1, encoding the active-zone protein RIMBP1, cause autosomal recessive dystonia. J Clin Invest. 2021;131. doi:10.1172/JCI140625

51. Suthar SK, Alam MM, Lee J, Monga J, Joseph A, Lee S-Y. Bioinformatic Analyses of Canonical Pathways of TSPOAP1 and its Roles in Human Diseases. Frontiers in Molecular Biosciences. 2021. Available: https://www.frontiersin.org/articles/10.3389/fmolb.2021.667947

52. Jun GR, Chung J, Mez J, Barber R, Beecham GW, Bennett DA, et al. Transethnic genome-wide scan identifies novel Alzheimer’s disease loci. Alzheimers Dement. 2017;13: 727–738. doi:10.1016/j.jalz.2016.12.012

53. Dib S, Pahnke J, Gosselet F. Role of ABCA7 in Human Health and in Alzheimer’s Disease. Int J Mol Sci. 2021;22. doi:10.3390/ijms22094603

54. Kaltschmidt B, Helweg LP, Greiner JFW, Kaltschmidt C. NF-κB in neurodegenerative diseases: Recent evidence from human genetics. Front Mol Neurosci. 2022;15: 954541. doi:10.3389/fnmol.2022.954541

55. Wightman DP, Savage JE, de Leeuw CA, Jansen IE, Posthuma D. Rare variant aggregation in 148,508 exomes identifies genes associated with proxy dementia. Sci Rep. 2023;13: 2179. doi:10.1038/s41598-023-29108-8

56. Bellenguez C, Charbonnier C, Grenier-Boley B, Quenez O, Le Guennec K, Nicolas G, et al. Contribution to Alzheimer’s disease risk of rare variants in TREM2, SORL1, and ABCA7 in 1779 cases and 1273 controls. Neurobiol Aging. 2017;59: 220.e1–220.e9. 10.1016/j.neurobiolaging.2017.07.001

57. Moorman SM, Carr K, Greenfield EA. Childhood socioeconomic status and genetic risk for poorer cognition in later life. Soc Sci Med. 2018;212: 219–226. 10.1016/j.socscimed.2018.07.025

58. Kulick ER, Elkind MS V, Boehme AK, Joyce NR, Schupf N, Kaufman JD, et al. Long-term exposure to ambient air pollution, APOE-ε4 status, and cognitive decline in a cohort of older adults in northern Manhattan. Environ Int. 2020;136: 105440. doi:10.1016/j.envint.2019.105440

59. Seifan A, Schelke M, Obeng-Aduasare Y, Isaacson R. Early Life Epidemiology of Alzheimer’s Disease--A Critical Review. Neuroepidemiology. 2015;45: 237–254. doi:10.1159/000439568

60. Reddy PH, Manczak M, Yin X, Grady MC, Mitchell A, Tonk S, et al. Protective Effects of Indian Spice Curcumin Against Amyloid-β in Alzheimer’s Disease. J Alzheimers Dis. 2018;61: 843–866. doi:10.3233/JAD-170512

61. Peters R, Ee N, Peters J, Booth A, Mudway I, Anstey KJ. Air Pollution and Dementia: A Systematic Review. J Alzheimers Dis. 2019;70: S145–S163. doi:10.3233/JAD-180631

62. Banerjee J, Jain U, Khobragade P, Weerman B, Hu P, Chien S, et al. Methodological considerations in designing and implementing the harmonized diagnostic assessment of dementia for longitudinal aging study in India (LASI-DAD). Biodemography Soc Biol. 2020;65: 189–213. doi:10.1080/19485565.2020.1730156

63. Sengupta D, Choudhury A, Basu A, Ramsay M. Population Stratification and Underrepresentation of Indian Subcontinent Genetic Diversity in the 1000 Genomes Project Dataset. Genome Biol Evol. 2016;8: 3460–3470. doi:10.1093/gbe/evw244

64. Nakatsuka N, Moorjani P, Rai N, Sarkar B, Tandon A, Patterson N, et al. The promise of discovering population-specific disease-associated genes in South Asia. Nat Genet. 2017;49: 1403–1407. doi:10.1038/ng.3917

65. Moorjani P, Thangaraj K, Patterson N, Lipson M, Loh P-R, Govindaraj P, et al. Genetic evidence for recent population mixture in India. Am J Hum Genet. 2013;93: 422–438. doi:10.1016/j.ajhg.2013.07.006

