## Supplemental Material for "Region-based analysis with functional annotation identifies genes associated with cognitive function in South Asians from India"

**Supplemental Methods**

Gene Selection

We selected a total of 84 genes from EA GWAS of AD (Bellenguez et al.^1^ and Wightman et al.^2^) as well as genes in the *APOE* region previously associated with AD (*TOMM40* and *APOC1*; Supplemental Table 1) .^2–5^ From Bellenguez et al.,^1^ we selected a total of 73 genes that were either identified as known loci (i.e., appeared in Bellenguez Table 1) or that had the strongest evidence of driving the detected novel AD associations (tier 1 in gene prioritization; see Bellenguez Supplementary Table 20). From Wightman et al.,^2^ we selected 45 genes from independent loci that reached genome-wide significance (see Wightman Table 1).

For any loci that were listed in the format “gene A/gene B”, both genes were included in our analysis, and we examined the gene region to identify additional genes that may be relevant within the locus. Five loci from Bellenguez Table 1 and four loci from Wightman Table 1 were listed as “gene A/gene B”, with an overlap of three loci between them. For the locus HLA from Bellenguez Table 1, we used *HLA-DQA1, HLA-DRB1*, and *HLA-DRB5* separately because this locus was labeled as *HLA-DRB1* and *HLA-DRB5* in other AD GWAS,^6–9^ and *HLA-DQA1* was labeled as the closest gene in Bellenguez et al.^1^ For the locus MS4A, we used the genes *MS4A4A* and *MS4A6A* separately, as MS4A has been labeled as *MS4A6A*,^7,8^ *MS4A2*,^6^ and *MS4A4A*^1^,^2^ in various AD GWAS. Finally, for *CELF1/SPI1*, we also used *CELF1, SPI1*, and *MADD* separately. This is due to the locus *CELF1/SPI1* being labeled as *CELF1*,^8^ *SPI1/CELF1*,^6^ and *MADD/SPI1*^9^ in various AD GWAS. The *ZCWPW1/NYAP1* locus has been labeled as *ZCWPW1*^7,8^ and *ZCWPW1/NYAP1*^6,9^. While *SPYDE3* was listed as the closest gene in Bellenguez^1^ to the *ZCWPW1/NYAP1* locus, we did not examine this gene because AD GWAS did not specify *SPYDE3* as a prioritized gene. The *SLC24A4/RIN3* locus has been labeled as *SLC24A4/RIN3,*^8^ *SLC24A4,*^6,7^ and *RIN3*^9^ in previous GWAS.

Annotation Scores

For missense/LoF SNVs, we used CADD_raw_rankscore, a measure of variant deleteriousness combining multiple genomic features of each variant;^10^ GERP_RS_rankscore, a measure of variant conservation;^11^ Eigen.phred, a measure of variant deleteriousness using an unsupervised learning method;^12^ and fathmm.MKL_coding_rankscore, a score from a machine learning method incorporating other annotations to predict deleteriousness of the variant from coding variants.^13^ For promoter/enhancer SNVs, we used CADD_raw_rankscore,^10^ GERP_RS_rankscore,^11^ Eigen.PC.phred,^12^ fathmm.MKL_non.coding_rankscore,^13^ and GenoCanyon_rankscore, a measure of variant conservation.^14^ The rank score variables are a number between zero and one indicating the genome-wide ranks of the corresponding annotation score across all variants as retrieved via WGS Annotator. Higher rank score variables indicate higher annotation scores. Per the STAAR tutorial, Phred scores were calculated from the rank scores according to the equation -10*log_10_(1-x), where x is the rank score. For Eigen.phred and Eigen.PC.phred, these Phred scores were provided from the original database using either coding or non-coding variant training data. Almost all missense/LoF SNVs had Eigen.phred calculated using the coding training data set, representing a Phred score among all coding variants. Almost all promoter/enhancer SNVs had Eigen.PC.phred calculated with a non-coding variant training dataset, representing a Phred score among all non-coding variants.

**Supplemental Table 1**: Sources used to identify the 84 genes analyzed in this study.

| Gene | Locus | Location in Bellenguez et al. (2022)* | Location in Wightman et al. (2021)* |
| --- | --- | --- | --- |
| *ABCA1* | *ABCA1* | Tier 1 | N/A |
| *ABCA7* | *ABCA7* | Table 1 | Table 1 |
| *ABI3* | *ABI3* | Table 1 | Table 1 |
| *ACE* | *ACE* | Table 1 | Table 1 |
| *ADAM10* | *ADAM10* | Table 1 | Table 1 |
| *ADAM17* | *ADAM17* | Tier 1 | N/A |
| *ADAMTS1* | *ADAMTS1* | Table 1 | N/A |
| *AGRN* | *AGRN* | N/A | Table 1 |
| *APH1B* | *APH1B* | Table 1 | Table 1 |
| *APOC1* | *APOE* | N/A | N/A |
| *APOE* | *APOE* | N/A | Table 1 |
| *APP* | *APP* | Tier 1 | Table 1 |
| *BIN1* | *BIN1* | Table 1 | Table 1 |
| *BLNK* | *BLNK* | Tier 1 | N/A |
| *CASS4* | *CASS4* | Table 1 | Table 1 |
| *CCDC6* | *ANK3* | Tier 1 | Table 1 |
| *CD2AP* | *CD2AP* | Table 1 | Table 1 |
| *CD33* | *CD33* | N/A | Table 1 |
| *CELF1* | *CELF1/SPI1* | Table 1 | N/A |
| *CLNK* | *CLNK/HS3ST1* | Table 1 | Table 1 |
| *CLU* | *CLU* | Table 1 | Table 1 |
| *CR1* | *CR1* | Table 1 | Table 1 |
| *CTSB* | *CTSB* | Tier 1 | N/A |
| *CTSH* | *CTSH* | Tier 1 | N/A |
| *DGKQ* | *IDUA* | Tier 1 | N/A |
| *DOC2A* | *DOC2A* | Tier 1 | N/A |
| *ECHDC3* | *USP6NL/ECHDC3* | Table 1 | Table 1 |
| *EGFR* | *SEC61G* | Tier 1 | N/A |
| *EPHA1* | *EPHA1/EPHA1-AS1* | Table 1 | N/A |
| *EPHA1-AS1* | *EPHA1/EPHA1-AS1* | N/A | Table 1 |
| *FERMT2* | *FERMT2* | Table 1 | Table 1 |
| *GRN* | *GRN* | Tier 1 | Table 1 |
| *HAVCR2* | *HAVCR2* | N/A | Table 1 |
| *HLA-DQA1* | *HLA* | Table 1 | Table 1 |
| *HLA-DRB1* | *HLA* | Table 1 | Table 1 |
| *HLA-DRB5* | *HLA* | Table 1 | Table 1 |
| *HS3ST1* | *CLNK/HS3ST1* | Table 1 | N/A |
| *ICA1* | *ICA1* | Tier 1 | N/A |
| *ICA1L* | *WDR12* | Tier 1 | N/A |
| *IL34* | *IL34* | Table 1 | N/A |
| *INPP5D* | *INPP5D* | Table 1 | Table 1 |
| *JAZF1* | *JAZF1* | Tier 1 | N/A |
| *KAT8* | *KAT8* | Table 1 | N/A |
| *LILRB2* | *LILRB2* | Tier 2 | Table 1 |
| *LIME1* | *SLC2A4RG* | Tier 1 | N/A |
| *MADD* | *CELF1/SPI1* | Table 1 | Table 1 |
| *MAF* | *MAF* | Tier 1 | N/A |
| *MAPT* | *MAPT* | Table 1 | N/A |
| *MME* | *MME* | Tier 1 | N/A |
| *MS4A4A* | *MS4A* | Table 1 | Table 1 |
| *MS4A6A* | *MS4A* | Table 1 | Table 1 |
| *MYO15A* | *MYO15A* | Tier 1 | N/A |
| *NCK2* | *NCK2* | Tier 1 | Table 1 |
| *NME8* | *NME8* | Table 1 | N/A |
| *NTN5* | *NTN5* | N/A | Table 1 |
| *NYAP1* | *ZCWPW1/NYAP1* | Table 1 | Table 1 |
| *OTULIN* | *ANKH* | Tier 1 | N/A |
| *PICALM* | *PICALM* | Table 1 | Table 1 |
| *PLCG2* | *PLCG2* | Table 1 | N/A |
| *PLEKHA1* | *PLEKHA1* | Tier 1 | N/A |
| *PTK2B* | *PTK2B* | Table 1 | N/A |
| *RABEP1* | *SCIMP/RABEP1* | Table 1 | Table 1 |
| *RBCK1* | *RBCK1* | Tier 1 | N/A |
| *RHOH* | *RHOH* | Tier 1 | N/A |
| *RIN3* | *SLC24A4/RIN3* | Table 1 | Table 1 |
| *RITA1* | *TPCN1* | Tier 1 | N/A |
| *SCIMP* | *SCIMP/RABEP1* | Table 1 | Table 1 |
| *SHARPIN* | *SHARPIN* | Tier 1 | Table 1 |
| *SIGLEC11* | *SIGLEC11* | Tier 1 | N/A |
| *SLC24A4* | *SLC24A4/RIN3* | Table 1 | N/A |
| *SORL1* | *SORL1* | Table 1 | Table 1 |
| *SORT1* | *SORT1* | Tier 1 | N/A |
| *SPI1* | *CELF1/SPI1* | Table 1 | Table 1 |
| *SPPL2A* | *SPPL2A* | Table 1 | N/A |
| *TMEM106B* | *TMEM106B* | Tier 1 | Table 1 |
| *TNIP1* | *TNIP1* | Tier 1 | Table 1 |
| *TOMM40* | *APOE* | N/A | N/A |
| *TREM2* | *TREM2* | Table 1 | Table 1 |
| *TSPAN14* | *TSPAN14* | Tier 1 | N/A |
| *TSPOAP1* | *TSPOAP1/TSPOAP1-AS1* | Table 1 | N/A |
| *TSPOAP1-AS1* | *TSPOAP1/TSPOAP1-AS1* | N/A | Table 1 |
| *USP6NL* | *USP6NL/ECHDC3* | N/A | Table 1 |
| *WDR81* | *WDR81* | Tier 1 | N/A |
| *ZCWPW1* | *ZCWPW1/NYAP1* | Table 1 | Table 1 |
| *Indicates the location or prioritization of each gene in the Bellenguez et al. (2022) or Wightman et al. (2021) genome-wide association study | | | |

**Supplemental Table 2:** Summary of cognitive measures in LASI-DAD

| **Phenotype** | **Minimum** | **Q1** | **Median** | **Q3** | **Maximum** | **Mean** | **SD** |
| --- | --- | --- | --- | --- | --- | --- | --- |
| HMSE score | 0 | 19 | 24 | 27 | 30 | 22.7 | 5.39 |
| General Cognitive Function | -3.03 | -0.68 | -0.044 | 0.68 | 2.77 | 0.010 | 0.92 |
| Memory | -2.20 | -0.63 | -0.049 | 0.64 | 3.63 | 0.022 | 0.94 |
| Executive Function | -1.93 | -0.69 | -0.067 | 0.65 | 2.48 | -0.001 | 0.90 |
| Orientation | -2.48 | -0.55 | -0.003 | 0.94 | 0.94 | -0.020 | 0.79 |
| Language/Fluency | -3.37 | -0.55 | 0.003 | 0.55 | 1.96 | -0.031 | 0.80 |
| Visuospatial | -1.58 | -0.73 | -0.104 | 0.54 | 1.58 | 0.036 | 0.83 |

HMSE = Hindi Mental State Exam, SD = Standard Deviation, Q1 = Quartile 1, Q3 = Quartile 3

**Supplemental Table 3** Annotation weight distribution for missense/LoF SNVs.

| **Annotation** | **Minimum** | **Q1** | **Median** | **Q3** | **Maximum** | **Mean** |
| --- | --- | --- | --- | --- | --- | --- |
| CADD_raw_rankscore | 0.00 | 0.95 | 0.99 | 1.00 | 1.00 | 0.87 |
| fathmm_MKL_coding_rankscore | 0.00 | 0.90 | 0.98 | 0.99 | 1.00 | 0.89 |
| GERP_RS_rankscore | 0.00 | 0.78 | 0.97 | 0.99 | 1.00 | 0.79 |
| Eigen-Phred | 0.00 | 0.70 | 2.21 | 4.75 | 27.05 | 3.25 |
| Transformed Eigen-Phred* | 0.00 | 0.15 | 0.40 | 0.67 | 1.00 | 0.42 |
| *Transformed by applying the equation 1 – 10^(-(Eigen-Phred)/10) to compare to rank score variables on a scale from 0 to 1.  Annotation weights taken from unique variants analyzed in our analysis,  Variants were analyzed if they had a minor allele frequency (MAF)>0 and a complete set of annotation weights. | | | | | | |

**Supplemental Table 4:** Annotation weight distribution for promoter/enhancer SNVs.

| **Annotation** | **Minimum** | **Q1** | **Median** | **Q3** | **Maximum** | **Mean** |
| --- | --- | --- | --- | --- | --- | --- |
| CADD_raw_rankscore | 0.00 | 0.44 | 0.76 | 0.91 | 1.00 | 0.66 |
| fathmm_MKL_non-coding_rankscore | 0.00 | 0.57 | 0.79 | 0.91 | 1.00 | 0.72 |
| GERP_RS_rankscore | 0.00 | 0.14 | 0.45 | 0.85 | 1.00 | 0.49 |
| GenoCanyon_score | 0.32 | 0.93 | 0.97 | 0.97 | 1.00 | 0.94 |
| Eigen-PC-Phred | 0.00 | 10.53 | 16.80 | 22.51 | 48.24 | 16.60 |
| Transformed Eigen-PC-Phred* | 0.00 | 0.91 | 0.98 | 0.99 | 1.00 | 0.90 |
| *Transformed by applying the equation 1 – 10^(-(Eigen-PC-Phred)/10) to compare to rank score variables on a scale from 0 to 1.  Annotation weights taken from unique variants analyzed in our analysis,  Variants were analyzed if they had a minor allele frequency (MAF)>0 and a complete set of annotation weights. | | | | | | |

**Supplemental Table 5:** Genes with at least one nominally significant association (p<0.05) in Missense/LoF Analysis (Model 1)

| **Gene** | **# of Variants Analyzed** | **P-value**  **(without annotation weights)** | **P-value**  **(with annotation weights)** |
| --- | --- | --- | --- |
| **HMSE Score** | | | |
| *ADAM17* | 15 | **0.017** | **0.021** |
| *APOE* | 20 | **9.5x10^-4^*** | **0.001*** |
| *PICALM* | 16 | **0.002*** | **0.002*** |
| *ABCA7* | 178 | 0.074 | **0.039** |
| *MS4A6A* | 16 | 0.063 | **0.049** |
| **General Cognitive Function** | | | |
| *OTULIN* | 15 | **0.013** | **0.028** |
| *APOE* | 20 | **5.6x10^-4^*** | **7.8x10^-4^*** |
| *LILRB2* | 69 | **0.024** | 0.066 |
| *TSPOAP1* | 89 | **0.006** | **0.013** |
| *ABCA7* | 178 | 0.051 | **0.034** |
| **Memory** | | | |
| *APOE* | 20 | **0.002** | **0.002** |
| *TSPOAP1* | 89 | **0.004** | **0.007** |
| *MAF* | 48 | 0.066 | **0.048** |
| **Executive Function** | | | |
| *ADAM17* | 15 | **0.029** | **0.036** |
| *APOE* | 20 | **0.002*** | **0.002** |
| *MAPT* | 46 | **0.045** | **0.047** |
| *TSPOAP1* | 89 | **0.002*** | **0.004** |
| **Orientation** |  |  |  |
| *ABCA7* | 178 | **0.018** | **0.008** |
| *CCDC6* | 9 | **0.022** | **0.022** |
| *OTULIN* | 15 | **0.025** | 0.056 |
| *APOE* | 20 | **9.3x10^-4^*** | **0.001** |
| *MAF* | 48 | **0.029** | **0.036** |
| *TSPOAP1* | 89 | **0.049** | 0.106 |
| *ABI3* | 18 | 0.061 | **0.031** |
| **Language/Fluency** | | | |
| *OTULIN* | 15 | **0.037** | 0.082 |
| *APOE* | 20 | **0.028** | **0.024** |
| *CTSH* | 17 | **0.019** | **0.029** |
| *NTN5* | 36 | **0.011** | **0.016** |
| *SORL1* | 96 | **0.022** | **0.031** |
| *ECHDC3* | 21 | **0.026** | **0.043** |
| **Visuospatial** | | | |
| *ADAM17* | 15 | **0.042** | 0.052 |
| *CR1* | 77 | **0.028** | **0.024** |
| *NCK2* | 8 | **0.009** | **0.010** |

HMSE = Hindi Mental State Exam, FDR = False Discovery Rate

Genes were included if the P-value with or without annotation weights was <0.05 in Model 1.

P-values<0.05 are in bold.

*FDR q-values<0.1

**Supplemental Table 6**: Genes with at least one nominally significant association (p<0.05) in Missense/LoF Analysis (Model 2)

| **Gene** | **# of Variants Analyzed** | **P-value**  **(without annotation weights)** | **P-value**  **(with annotation weights)** |
| --- | --- | --- | --- |
| **HMSE Score** | | | |
| *ABCA1* | 79 | **0.017** | **0.017** |
| *APOE* | 20 | **0.010** | **0.016** |
| *PICALM* | 16 | **0.001*** | **0.001*** |
| *MS4A6A* | 15 | 0.059 | **0.045** |
| **General Cognitive Function** | | | |
| *ABCA7* | 178 | **0.016** | **0.001** |
| *CCDC6* | 9 | **0.042** | **0.042** |
| *APOE* | 20 | **0.027** | **0.039** |
| *MME* | 31 | **0.026** | **0.027** |
| *TSPOAP1* | 89 | **0.017** | **0.024** |
| *HLA-DRB5* | 16 | 0.070 | **0.050** |
| **Memory** | | | |
| *CASS4* | 37 | **0.030** | **0.029** |
| *TSPOAP1* | 89 | **0.014** | **0.022** |
| *APOE* | 20 | **0.030** | **0.044** |
| *BLNK* | 12 | 0.053 | **0.042** |
| **Executive Function** | | | |
| *TSPOAP1* | 89 | **0.011** | **0.015** |
| *CCDC6* | 9 | **0.020** | **0.021** |
| *EPHA1-AS1* | 31 | **0.033** | **0.034** |
| **Orientation** | | | |
| *ABCA7* | 178 | **0.009** | **0.005** |
| *ABI3* | 18 | **0.024** | **0.013** |
| *CCDC6* | 9 | **0.008** | **0.008** |
| *APOE* | 20 | **0.006** | **0.009** |
| *DGKQ* | 38 | **0.029** | **0.033** |
| *RBCK1* | 17 | **0.034** | **0.035** |
| *HLA-DRB5* | 16 | 0.064 | **0.047** |
| **Language/Fluency** | | | |
| *TSPOAP1* | 89 | **0.050** | **0.031** |
| *MME* | 31 | **0.048** | **0.049** |
| *TREM2* | 11 | **0.040** | **0.039** |
| *PLEKHA1* | 19 | **0.035** | **0.036** |
| *CTSH* | 17 | **0.014** | **0.021** |
| *SORL1* | 96 | **0.006** | **0.006** |
| **Visuospatial** | | | |
| *ABCA1* | 79 | **0.042** | **0.031** |
| *ADAM10* | 14 | **0.025** | 1 |
| *ADAM17* | 15 | **0.046** | 0.057 |
| *APOE* | 20 | **0.028** | **0.022** |
| *NCK2* | 8 | **0.034** | **0.038** |
| *PLEKHA1* | 19 | **0.014** | **0.015** |
| *NTN5* | 36 | 0.067 | **0.034** |

HMSE = Hindi Mental State Exam, FDR = False Discovery Rate

Genes were included if the P-value with or without annotation weights was <0.05 in Model 2.

P-values<0.05 are in bold.

*FDR q-values<0.1.

**Supplemental Table 7:** Genes with at least one nominally significant (p<0.05) association in the high-confidence missense/LoF SNV analysis in Model 1.

| **Gene** | **# Of Variants Analyzed** | **P-Value (No Annotation Weights)** | **P-Value (Annotation Weights)** |
| --- | --- | --- | --- |
| **General Cognitive Function** |  |  |  |
| *DGKQ* | 2 | **0.042** | **0.042** |
| *INPP5D* | 8 | **0.028** | **0.026** |
| **Memory** |  |  |  |
| *MAPT* | 2 | **0.025** | **0.025** |
| **Executive Function** |  |  |  |
| *ADAMTS1* | 13 | **0.028** | **0.030** |
| *DGKQ* | 2 | **2.1x10^-3^** | **2.1x10^-3^** |
| *INPP5D* | 8 | **0.038** | **0.035** |
| **Orientation** |  |  |  |
| *ABCA7* | 63 | **0.016** | **0.021** |
| *ABI3* | 3 | **0.044** | **0.045** |
| **Language/Fluency** |  |  |  |
| *APOE* | 7 | **0.018** | **0.018** |
| **Visuospatial** |  |  |  |
| *ICA1* | 11 | **0.026** | **0.024** |
| *INPP5D* | 8 | **0.022** | **0.020** |
| HMSE = Hindi Mental State Exam  Genes were included if the P-value with or without annotation weights was less than 0.05 Bolded values are p<0.05. | | | |

**Supplemental Table 8:** Genes with at least one nominally significant (p<0.05) association in the high-confidence missense/LoF SNV analysis in Model 2

| **Gene** | **# Of Variants Analyzed** | **P-Value (No Annotation Weights)** | **P-Value (Annotation Weights)** |
| --- | --- | --- | --- |
| **Executive Function** | | | |
| *DGKQ* | 2 | **2.9x10^-3^** | **2.8x10^-3^** |
| *EPHA1-AS1* | 3 | **0.029** | **0.032** |
| *INPP5D* | 8 | 0.051 | **0.048** |
| **Orientation** | | | |
| *ABI3* | 3 | **0.039** | **0.040** |
| *ADAM17* | 3 | **0.034** | **0.031** |
| *SLC24A4* | 11 | **0.048** | **0.043** |
| **Language/Fluency** | | | |
| *EPHA1-AS1* | 3 | **0.033** | **0.042** |
| **Visuospatial** | | | |
| *ABCA1* | 36 | **0.035** | **0.039** |
| *APOE* | 7 | **0.015** | **0.015** |
| *GRN* | 10 | **0.025** | **0.025** |
| *INPP5D* | 8 | **0.041** | **0.039** |
| *RABEP1* | 2 | **5.2x10^-3^** | **5.3x10^-3^** |
| HMSE = Hindi Mental State Exam  Genes were included if the P-value with or without annotation weights was less than 0.05 Bolded values are p<0.05. | | | |

**Supplemental Table 9:** Nominally associated genes in the Brain-Specific Promoter/Enhancer analysis in Model 1.

| **Gene** | **# of Variants Analyzed** | **P-value**  **(without annotation weights)** | **P-value**  **(with annotation weights)** |
| --- | --- | --- | --- |
| **HMSE Score** | | | |
| *APOE* | 101 | **0.017** | **0.018** |
| *CCDC6* | 125 | **0.017** | **0.021** |
| *PLCG2* | 146 | **0.046** | **0.045** |
| *SCIMP* | 28 | **0.037** | **0.019** |
| **General Cognitive Function** | | | |
| *APOE* | 101 | **0.008** | **0.009** |
| *BLNK* | 98 | **0.044** | **0.038** |
| *KAT8* | 52 | **0.040** | 0.055 |
| *SCIMP* | 28 | **0.022** | **0.026** |
| *TSPOAP1* | 231 | **0.020** | **0.021** |
| *TSPOAP1-AS1* | 158 | **0.009** | **0.011** |
| *APOC1* | 93 | 0.060 | **0.050** |
| **Memory** | | | |
| *APOC1* | 93 | **0.037** | **0.029** |
| *APOE* | 101 | **0.026** | **0.028** |
| *INPP5D* | 168 | **0.047** | 0.066 |
| *JAZF1* | 71 | **0.025** | **0.027** |
| *SCIMP* | 28 | **0.006** | **0.008** |
| *TSPOAP1* | 231 | **0.006** | **0.006** |
| *TSPOAP1-AS1* | 158 | **0.003** | **0.004** |
| **Executive Function** | | | |
| *APOE* | 101 | **0.025** | **0.027** |
| *BIN1* | 213 | **0.013** | **0.012** |
| *CD33* | 23 | **0.040** | **0.016** |
| *MAPT* | 151 | **0.026** | **0.026** |
| *SHARPIN* | 116 | **0.033** | **0.031** |
| *TMEM106B* | 129 | **0.020** | **0.020** |
| *TSPOAP1* | 231 | **0.019** | **0.022** |
| *TSPOAP1-AS1* | 158 | **0.012** | **0.015** |
| **Orientation** | | | |
| *ADAM17* | 80 | **0.027** | **0.022** |
| *APOE* | 101 | **0.015** | **0.017** |
| *SCIMP* | 28 | **0.023** | **0.026** |
| *USP6NL* | 100 | **0.037** | 0.083 |
| **Language/Fluency** | | | |
| *CTSH* | 65 | **0.012** | **0.011** |
| *HLA-DQA1* | 26 | **0.049** | **0.043** |
| *HLA-DRB1* | 44 | **0.022** | **0.024** |
| *PLCG2* | 146 | **0.009** | **0.009** |
| *SCIMP* | 28 | **0.016** | **0.017** |
| **Visuospatial** | | | |
| *BLNK* | 98 | **0.046** | **0.028** |

HMSE = Hindi Mental State Exam, FDR = False Discovery Rate

Genes were included if the P-value with or without annotation weights was <0.05 in Model 1.

P-values<0.05 are in bold.

*FDR q-values<0.1.

**Supplemental Table 10:** Nominally associated genes in Brain-Specific Promoter/Enhancer analysis in Model 2.

| **Gene** | **# of Variants** | **P-value**  **(without annotation weights)** | **P-value**  **(with annotation weights)** |
| --- | --- | --- | --- |
| **HMSE Score** | | | |
| *BLNK* | 98 | **0.039** | **0.036** |
| *ADAM17* | 80 | 0.052 | **0.044** |
| **General Cognitive Function** | | | |
| *ADAM10* | 133 | **0.040** | **0.038** |
| *ADAM17* | 80 | **0.026** | **0.020** |
| *BLNK* | 98 | **0.007** | **0.006** |
| *FERMT2* | 132 | **0.036** | **0.045** |
| *TMEM106B* | 129 | **0.023** | **0.023** |
| **Memory** | | | |
| *ADAM10* | 133 | **0.049** | 0.095 |
| *JAZF1* | 71 | **0.012** | **0.013** |
| *SCIMP* | 28 | **0.008** | **0.012** |
| *TSPOAP1* | 231 | **0.019** | **0.021** |
| *TSPOAP1-AS1* | 158 | **0.008** | **0.011** |
| *APOC1* | 93 | 0.053 | **0.047** |
| **Executive Function** | | | |
| *ADAM17* | 80 | **0.036** | **0.028** |
| *BLNK* | 98 | **0.029** | **0.024** |
| *CD2AP* | 81 | **0.034** | **0.032** |
| *MAPT* | 151 | **0.042** | **0.043** |
| *RHOH* | 6 | **0.010** | **0.014** |
| *TMEM106B* | 129 | **0.042** | 0.051 |
| *TSPOAP1-AS1* | 158 | **0.046** | 0.055 |
| **Orientation** | | | |
| *ADAM17* | 80 | **0.010** | **0.007** |
| *BLNK* | 98 | **0.049** | **0.045** |
| *EGFR* | 127 | **0.049** | 0.064 |
| *FERMT2* | 132 | **0.018** | **0.023** |
| *MAF* | 225 | **0.024** | **0.033** |
| *TOMM40* | 100 | **0.044** | **0.035** |
| *APOC1* | 93 | 0.060 | **0.033** |
| **Language/Fluency** | | | |
| *CTSH* | 65 | **0.006** | **0.006** |
| *FERMT2* | 132 | **0.004** | **0.005** |
| *HLA-DQA1* | 26 | **0.031** | **0.023** |
| *INPP5D* | 168 | **0.029** | **0.043** |
| *PLCG2* | 146 | **0.012** | **0.012** |
| *SORT1* | 200 | 0.96 | **0.036** |
| **Visuospatial** | | | |
| *APOE* | 101 | **0.048** | 0.053 |
| *CD2AP* | 81 | **0.015** | **0.014** |
| *CR1* | 36 | **0.030** | **0.042** |
| *MS4A6A* | 59 | **0.048** | **0.048** |
| *TMEM106B* | 129 | **0.032** | **0.028** |

HMSE = Hindi Mental State Exam, FDR = False Discovery Rate

Genes were included if the P-value with or without annotation weights was <0.05 in Model 2.

P-values<0.05 are in bold.

*FDR q-values<0.1.

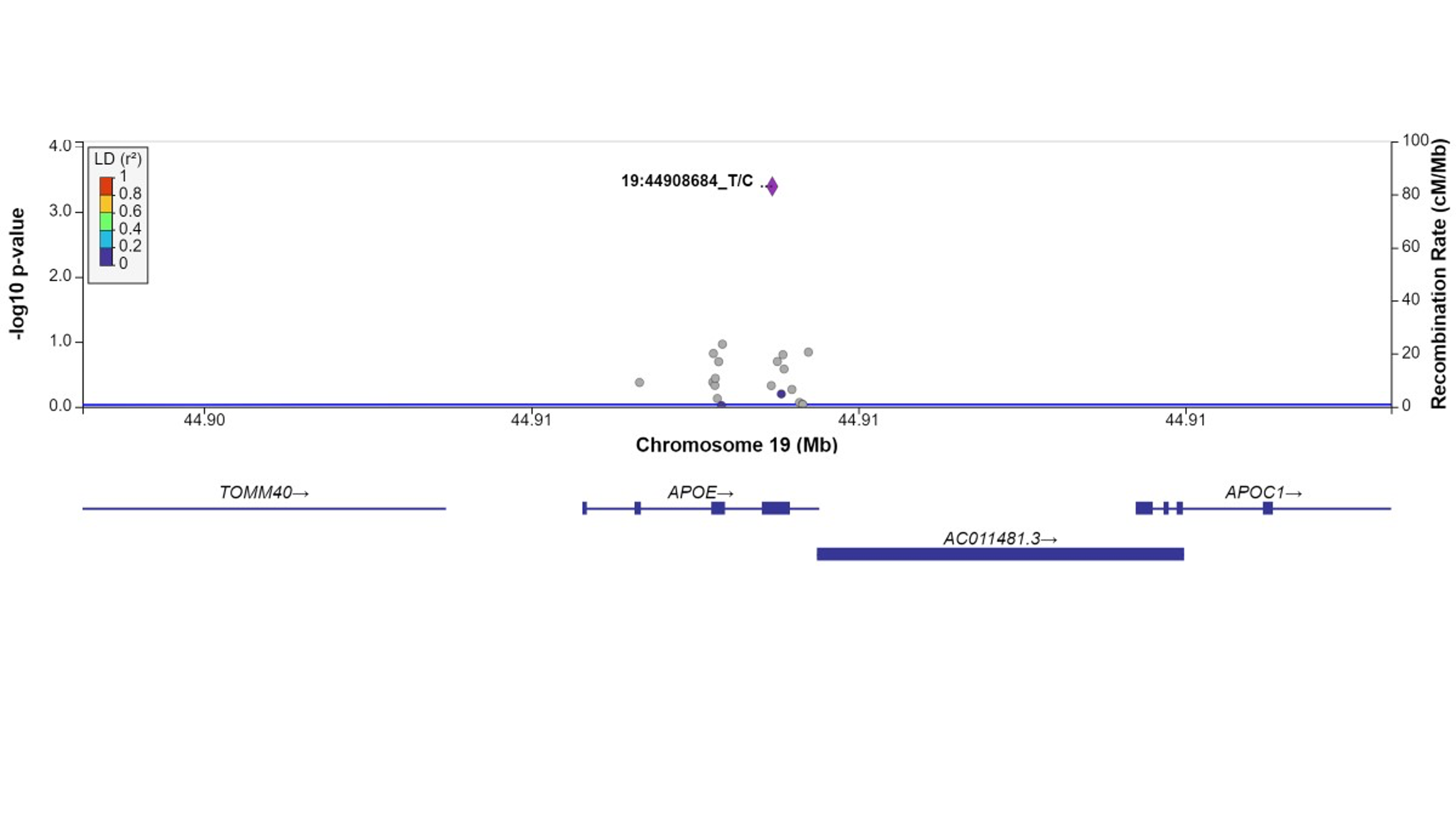

Supplemental Figure 1: Plot of missense/loss-of-function SNVs in APOE gene in Model 1 for executive function. Left Y-axis: -log10(p-value) from association between SNV and executive function, adjusting for age, sex, state/territory, the first 10 principal components of genetic ancestry; and accounting for relatedness and heteroscedastic variances among state/territory; Right Y-axis: SNV recombination rate based on HapMap GRCh38 SAS; X-axis: chromosomal location and gene regions; r^2^ color code: degree of linkage disequilibrium with index (most strongly associated) SNV, rs429358 (purple diamond). Grey points indicate no LD information present in the reference panel. No annotation weights were used to generate p-value.

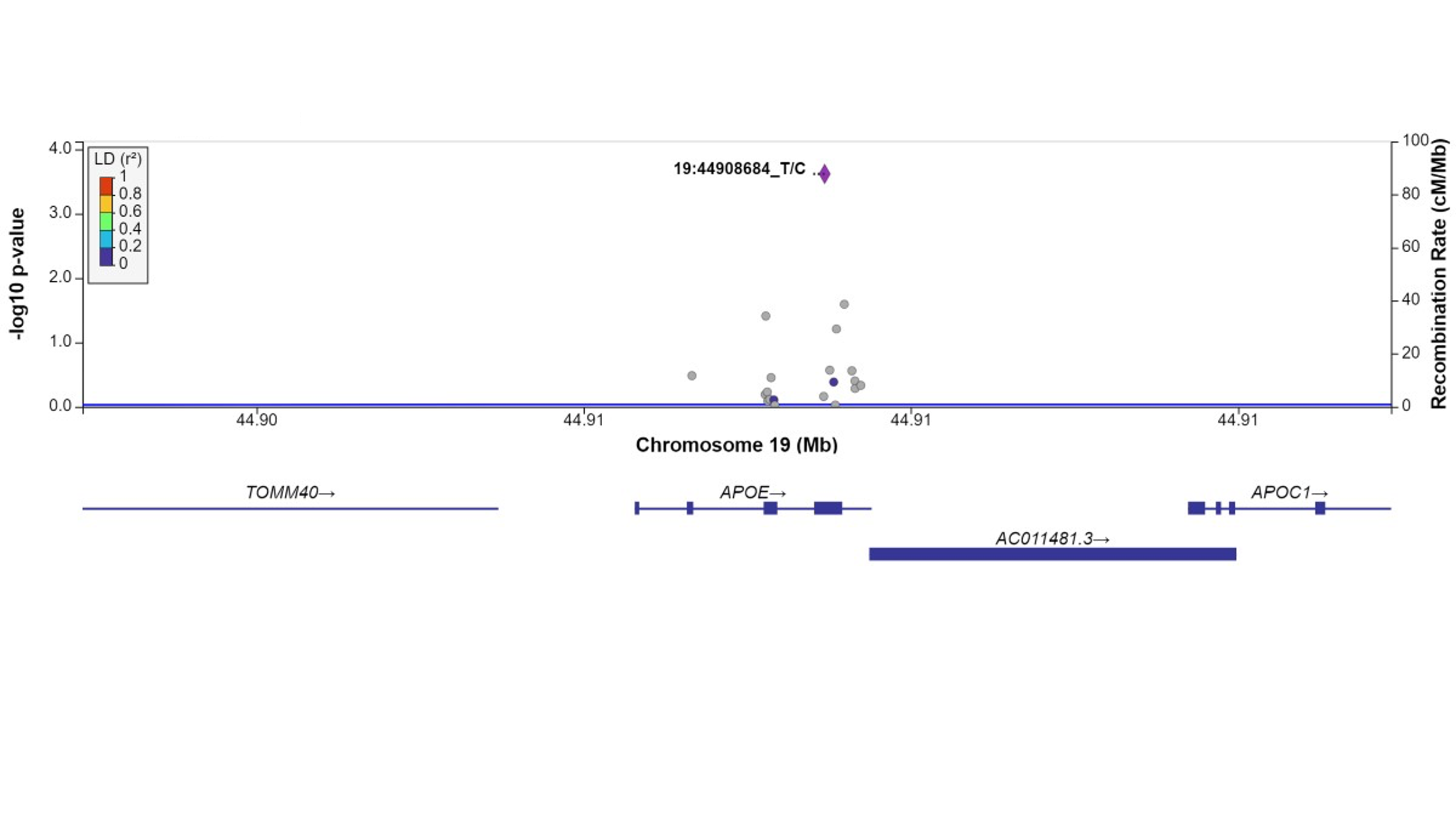

Supplemental Figure 2: Plot of missense/loss-of-function SNVs in APOE gene in Model 1 for orientation. Left Y-axis: -log10(p-value) from association between SNV and orientation, adjusting for age, sex, state/territory, the first 10 principal components of genetic ancestry; and accounting for relatedness and heteroscedastic variances among state/territory; Right Y-axis: SNV recombination rate based on HapMap GRCh38 SAS; X-axis: chromosomal location and gene regions; r^2^ color code: degree of linkage disequilibrium with index (most strongly associated) SNV, rs429358 (purple diamond). Grey points indicate no LD information present in the reference panel. No annotation weights were used to generate p-value.

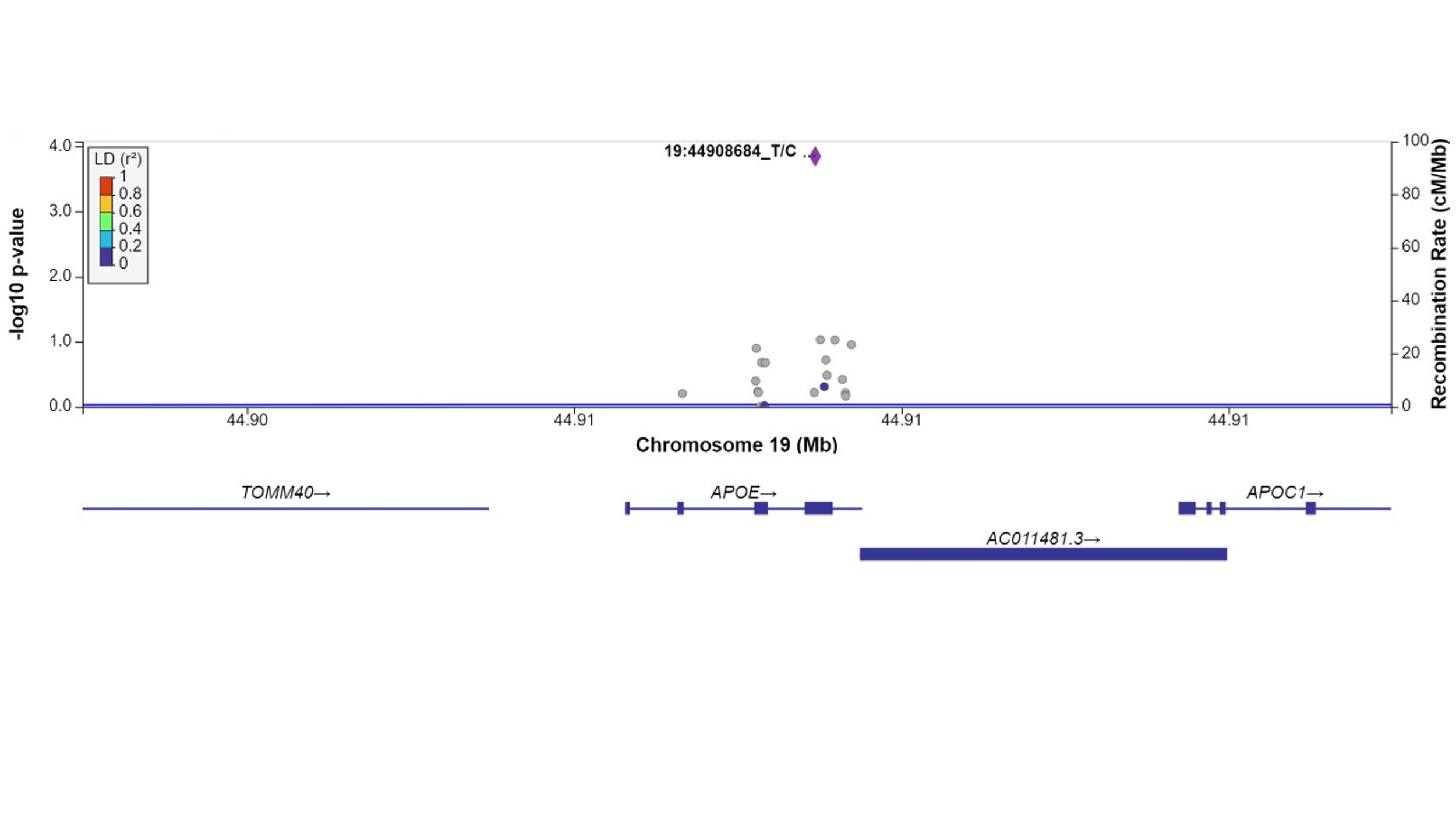

Supplemental Figure 3: Plot of missense/loss-of-function SNVs in APOE gene in Model 1 for general cognitive function. Left Y-axis: -log10(p-value) from association between SNV and general cognitive function, adjusting for age, sex, state/territory, the first 10 principal components of genetic ancestry; and accounting for relatedness and heteroscedastic variances among state/territory; Right Y-axis: SNV recombination rate based on HapMap GRCh38 SAS; X-axis: chromosomal location and gene regions; r^2^ color code: degree of linkage disequilibrium with index (most strongly associated) SNV, rs429358 (purple diamond). Grey points indicate no LD information present in the reference panel. No annotation weights were used to generate p-value.

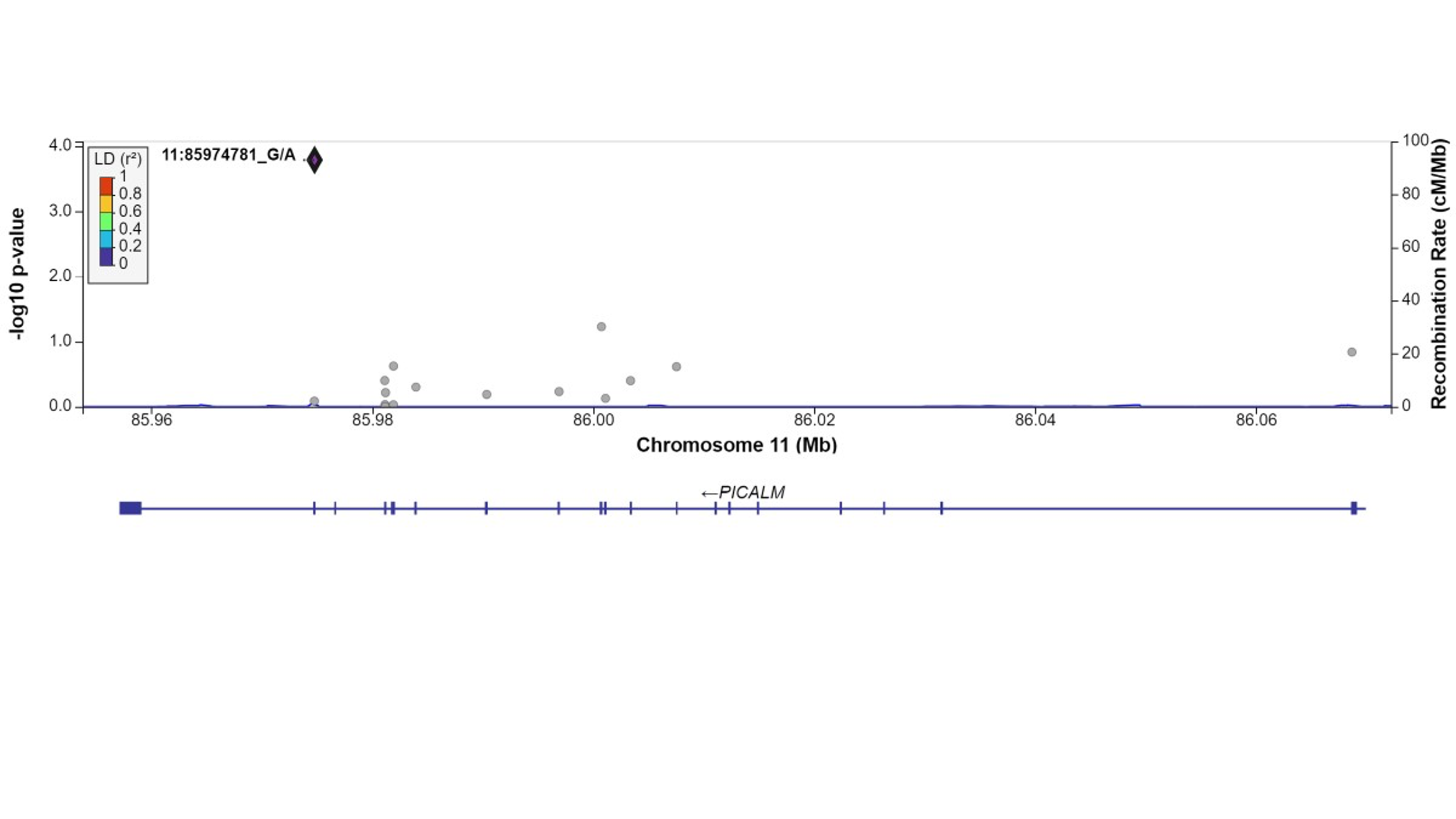

Supplemental Figure 4: Plot of missense/loss-of-function SNVs in PICALM gene in Model 2 for Hindi Mental State Exam (HMSE) score. Left Y-axis: -log10(p-value) from association between SNV and HMSE score, adjusting for age, sex, state/territory, education, literacy, urban/rural status, the first 10 principal components of genetic ancestry; and accounting for relatedness and heteroscedastic variances among state/territory; Right Y-axis: SNV recombination rate based on HapMap GRCh38 SAS; X-axis: chromosomal location and gene regions; r^2^ color code: degree of linkage disequilibrium with index (most strongly associated) SNV, rs779406084 (purple diamond). Grey points indicate no LD information present in the reference panel. No annotation weights were used to generate p-values.

**References**

1.	Bellenguez C, Küçükali F, Jansen IE, et al. New insights into the genetic etiology of Alzheimer’s disease and related dementias. *Nat Genet*. 2022;54(4):412-436. doi:10.1038/s41588-022-01024-z

2.	Zhou Q, Zhao F, Lv ZP, et al. Association between APOC1 polymorphism and alzheimer’s disease: A case-control study and meta-analysis. *PLoS One*. 2014;9(1). doi:10.1371/journal.pone.0087017

3.	Corder EH, Saunders AM, Strittmatter WJ, et al. Gene dose of apolipoprotein E type 4 allele and the risk of Alzheimer’s disease in late onset families. *Science (80- )*. 1993;261(5123):921-923. doi:10.1126/science.8346443

4.	Lutz MW, Crenshaw D, Welsh-Bohmer KA, Burns DK, Roses AD. New Genetic Approaches to AD: Lessons from APOE-TOMM40 Phylogenetics. *Curr Neurol Neurosci Rep*. 2016;16(5):48. doi:10.1007/s11910-016-0643-8

5.	Lin R, Zhang Y, Yan D, et al. Association of common variants in TOMM40/APOE/APOC1 region with human longevity in a Chinese population. *J Hum Genet*. 2016;61(4):323-328. doi:10.1038/jhg.2015.150

6.	Kunkle BW, Grenier-Boley B, Sims R, et al. Genetic meta-analysis of diagnosed Alzheimer’s disease identifies new risk loci and implicates Aβ, tau, immunity and lipid processing. *Nat Genet*. 2019;51(3):414-430. doi:10.1038/s41588-019-0358-2

7.	Jansen IE, Savage JE, Watanabe K, et al. Genome-wide meta-analysis identifies new loci and functional pathways influencing Alzheimer’s disease risk. *Nat Genet*. 2019;51(3):404-413. doi:10.1038/s41588-018-0311-9

8.	Lambert JC, Ibrahim-Verbaas CA, Harold D, et al. Meta-analysis of 74,046 individuals identifies 11 new susceptibility loci for Alzheimer’s disease. *Nat Genet*. 2013;45(12):1452-1458. doi:10.1038/ng.2802

9.	Wightman DP, Jansen IE, Savage JE, et al. A genome-wide association study with 1,126,563 individuals identifies new risk loci for Alzheimer’s disease. *Nat Genet*. 2021;53(9):1276-1282. doi:10.1038/s41588-021-00921-z

10.	Rentzsch P, Witten D, Cooper GM, Shendure J, Kircher M. CADD: Predicting the deleteriousness of variants throughout the human genome. *Nucleic Acids Res*. 2019;47(D1):D886-D894. doi:10.1093/nar/gky1016

11.	Davydov E V., Goode DL, Sirota M, Cooper GM, Sidow A, Batzoglou S. Identifying a high fraction of the human genome to be under selective constraint using GERP++. *PLoS Comput Biol*. 2010;6(12). doi:10.1371/journal.pcbi.1001025

12.	Ionita-Laza I, Mccallum K, Buxbaum J. A SPECTRAL APPROACH INTEGRATING FUNCTIONAL GENOMIC ANNOTATIONS FOR CODING AND NONCODING VARIANTS IULIANA IONITA-LAZA HHS Public Access Author manuscript. *Nat Genet*. 2016;48(2):214-220. doi:10.1038/ng.3477.A

13.	Shihab HA, Rogers MF, Gough J, et al. An integrative approach to predicting the functional effects of non-coding and coding sequence variation. *Bioinformatics*. 2015;31(10):1536-1543. doi:10.1093/bioinformatics/btv009

14.	Lu Q, Hu Y, Sun J, Cheng Y, Cheung KH, Zhao H. A statistical framework to predict functional non-coding regions in the human genome through integrated analysis of annotation data. *Sci Rep*. 2015;5(May):1-13. doi:10.1038/srep10576
